# Differences and similarities in diagnostic methods and treatments for Coronavirus disease 2019 (COVID-19): a scoping review

**DOI:** 10.1101/2020.10.30.20222950

**Authors:** Alessandro Rolim Scholze, Emiliana Cristina Melo, Carina Bortolato Major, Carolina Fordellone Rosa Cruz, Léia Regina de Souza Alcântara, Camila Dalcol, Fábio Rodrigues Ferreira Seiva, Maria de Fátima Mantovani, Ângela Tais Mattei, Henrique Spaulonci Silveira, Luiz Gustavo de Almeida Chuffa

## Abstract

**Aims:** We investigate a range of studies related to COVID-19 with focus on scientific evidence reporting the main diagnosis and treatments of the disease.

**Main Methods:** Scoping review conducted in the databases, MEDLINE, Cochrane, Embase, LILACS, Scopus, and Web of Science, and the gray Google Scholar literature, until May 2020. We follow PRISMA-SCR and the recommendations of the Joanna Briggs Institute. The identified studies were independently selected by peers. The qualitative data extracted were synthesized and organized into categories, and the quantitative data were generated through descriptive and inferential statistics.

**Key-findings:** 6060 articles were identified, of which 30 were included in this review. The publications are predominantly from China (n=22, 73.3%), and with a type of cross-sectional study (n=12, 40.0%), followed by a cohort (n=7, 23.0%). Among them, 16 studies addressed the diagnosis, and computed tomography was considered as non-invasive complementary method for detecting and evaluating the progression of COVID-19. Laboratory tests have been used to detect enzymatic or viral activities, and to monitor the inflammation associated with COVID-19. 14 studies included different therapeutic associations, such as Lopinavir/Ritonavir (LPV/r) and Arbidol, Hydroxychloroquine, Azithromycin, Tocilizumab and Remdesivir, and Corticosteroids/Plasminogen.

**Significance:** The evidence related to diagnostic methods are clear, and include tomography and laboratory tests. Medicinal or associated medications for the treatment of COVID-19, although showing a reduction in signs and COVID-19-related symptoms, can cause adverse effects of mild or severe intensity depending on viral load and inflammatory activity. Additional studies should be performed to identify the most reliable treatment for COVID-19.

## 1. Introduction

The current pandemic experienced globally, beginning in the city of Wuhan, China, at the end of 2019, is caused by a novel coronavirus called SARS-CoV-2, the disease being named COVID-19. Coronavirus is a family of viruses that cause respiratory diseases in humans, from the common cold to diseases such as Severe Acute Respiratory Syndrome (SARS) and Middle East Respiratory Syndrome (MERS), which resulted in high mortality rates in 2003y and 2012y, respectively [1]. To date, the disease has affected more than 27 million individuals in 216 countries, areas, or territories, culminating in more than 890 thousand confirmed deaths throughout the world. According to the number of cases, the most affected regions are the regions of the Americas, Southeast Asia, and Europe [1].

The main signs and symptoms of COVID-19 are multiple and include fever (83% -99%), cough (59% -82%), fatigue (44% -70%), decreased appetite (40% -84%), dyspnoea (31% -40%), myalgia (11% -35%), as well as other less specific symptoms involving the sore throat, nasal congestion, headache, diarrhea, nausea and vomiting, loss of olfactory sensitivity or taste [2]. In addition to the signs and symptoms, the diagnosis of COVID-19 are often performed by two methods: the molecular test and the serological test. The molecular test, known as RT-PCR, identifies the presence of the virus during the acute phase of the disease, and the serological test, also known as the rapid test, checks the antibody response of a given individual after days to weeks, indicating that the person has already been infected with SARS-CoV2. However, there are still uncertainties regarding the tests currently used with regard to sensitivity, specificity, and the ability to assess cross-reactivity with other types of coronavirus, such as SARS-CoV and MERS [1].

Regarding the available treatments, the agentes or compounds most often mentioned as “possible healing agents” are chloroquine, hydroxychloroquine, and azithromycin; however, the evidence is insufficient to indicate the use of a particular drug, alone or combined, during the treatment of COVID-19, and its use may be associated with more adverse effects than benefits. Following the incessant rush in searching for an effective treatment, several studies have adopted dubious measures in relation to the scientific robustness, in order to justify their clinical use [2].

## 2. Methods

We performed a scoping review to map the existing evidence in the literature on a given topic, published from different designs and study methodologies, as recommended by Aromataris [3]; in this review, we look at the diagnosis and treatment of COVID-19. This review followed five of six stages described in the framework of Aromataris [3], which was subsequently improved by Levac and collaborators [4], and recommended by Strumillo [5], namely: identifying the research question; identifying relevant studies; study selection; charting the data; collating, summarizing and reporting the results, and consultation (optional). The sixth stage of consulting the framework was not carried out.

### 2.1 Identifying the research question

The research questions were constructed from the elements of the PCC - Population (adults and elderly), Concept (diagnosis and treatment), and Context (COVID-19). The review questions explored in this study were: What types of diagnostic tests and drug treatments are available for adults and the elderly with COVID-19?

### 2.2 Identifying relevant studies

A systematic search was carried out between 14 and 15 May 2020, in seven electronic databases: MEDLINE (access via PubMed), Cochrane, Embase, Cumulative Index to Nursing and Allied Health Literature (CINAHL), Latin American Bibliographic Information (LILACS), Scopus and Web of Science (WoS), and in gray literature: Google Scholar. The search strategies adopted the terms: “diagnostic”, “treatment”, “laboratory techniques”, “COVID-19” and “SARS-CoV-2”, which were combined by Boolean operators “AND” and “OR” and were adapted according to each database. The filters used were: literature with human beings, in English, Spanish or Portuguese, complete articles, and published between the period from December 1, 2019, to May 15, 2020. **Table 1** illustrates the complete search strategy carried out at PUBMED.

**Table 1.** Search strategy performed using the PubMed database.

| Strategy | Descriptors | Results | Date/time |
| --- | --- | --- | --- |
| #1 | ("Adult"[Title/Abstract]) OR ("Aged"[Title/Abstract])<br>OR ("Elderly"[Title/Abstract]) OR ("Middle<br>aged"[Title/Abstract]) OR ("Ged over"[Title/Abstract])<br>AND ("Diagnosis"[Title/Abstract]) OR ("Clinical<br>laboratory techniques"[Title/Abstract]) OR ("Diagnostic<br>imaging"[Title/Abstract]) OR<br>("Radiography"[Title/Abstract]) OR ("COVID-19<br>diagnostic testing"[Title/Abstract]) OR ("COVID-19 drug<br>treatment"[Title/Abstract]) OR ("COVID-19<br>serotherapy"[Title/Abstract]) OR<br>("Therapeutics"[Title/Abstract]) OR<br>("Therapy"[Title/Abstract]) OR ("Drug<br>therapy"[Title/Abstract]) OR ("Duration of<br>therapy"[Title/Abstract]) OR ("Treatment<br>outcome"[Title/Abstract]) OR ("Clinical<br>protocols"[Title/Abstract]) AND<br>("Coronavirus"[Title/Abstract]) OR ("Coronavirus<br>Infections"[Title/Abstract]) OR ("COVID-19"[Title/Abstract]) OR ("SARS Virus"[Title/Abstract]) | 2.047 | 14/05/2020 - 23:36:25 |

**Table 1.**
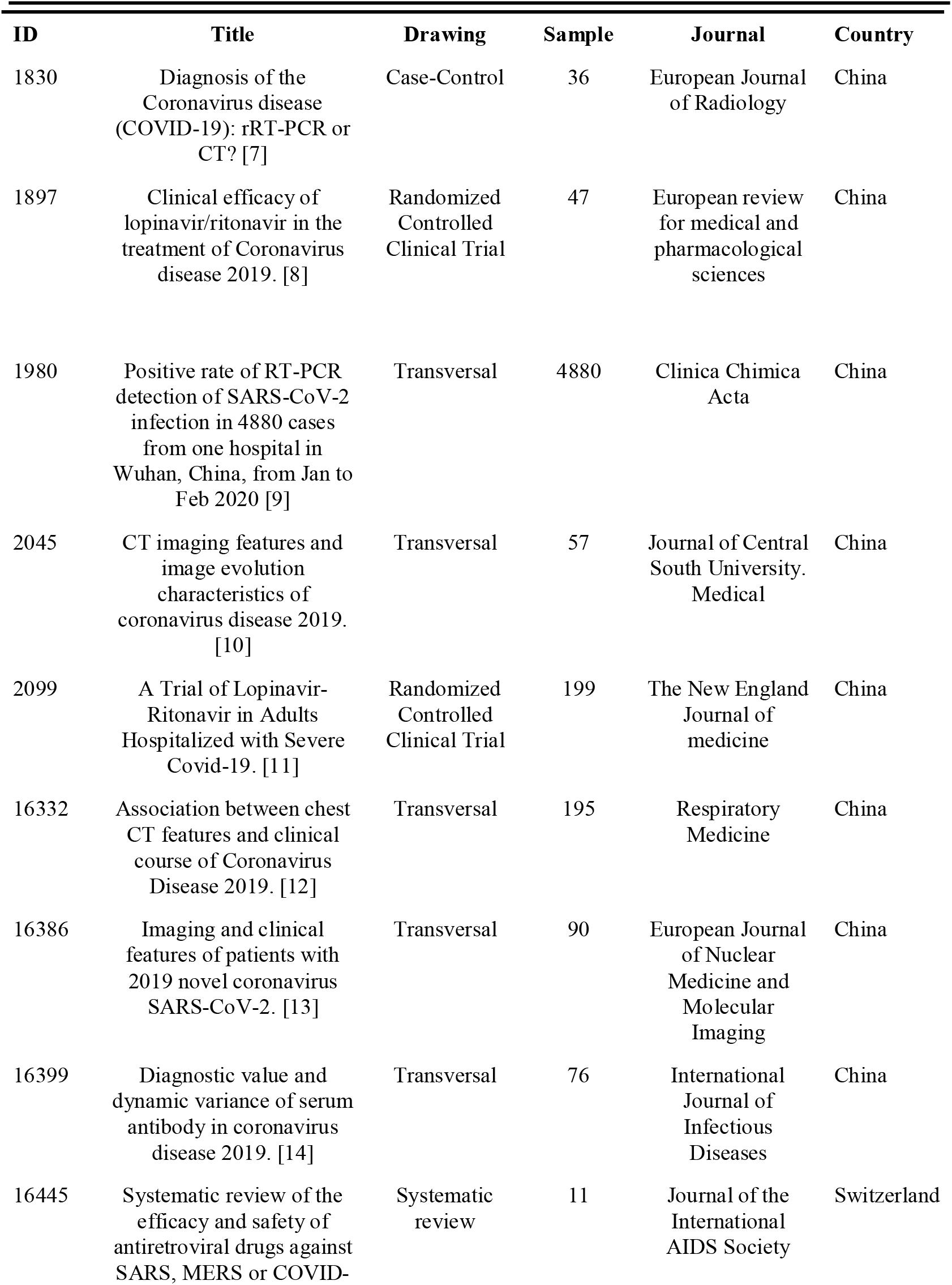
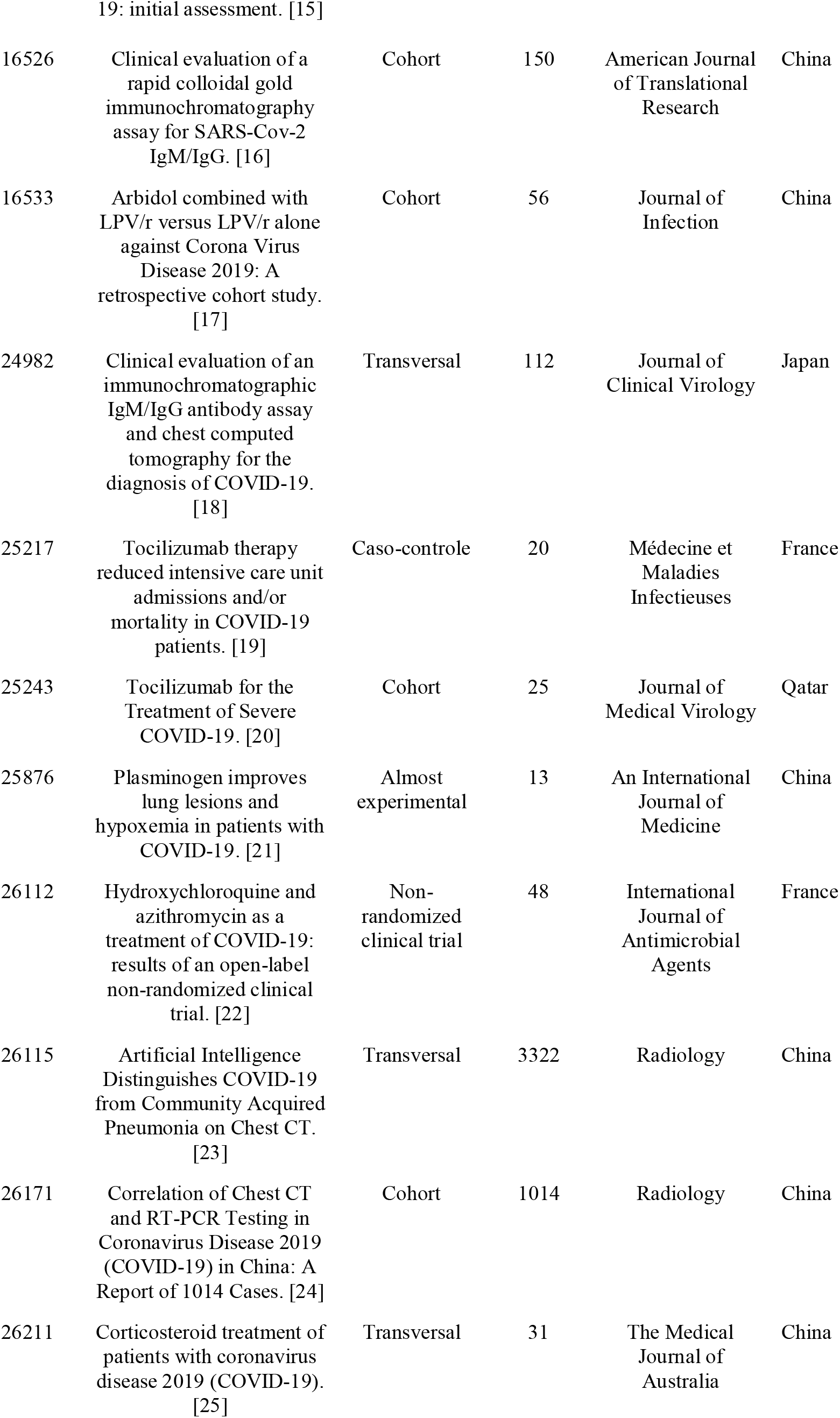
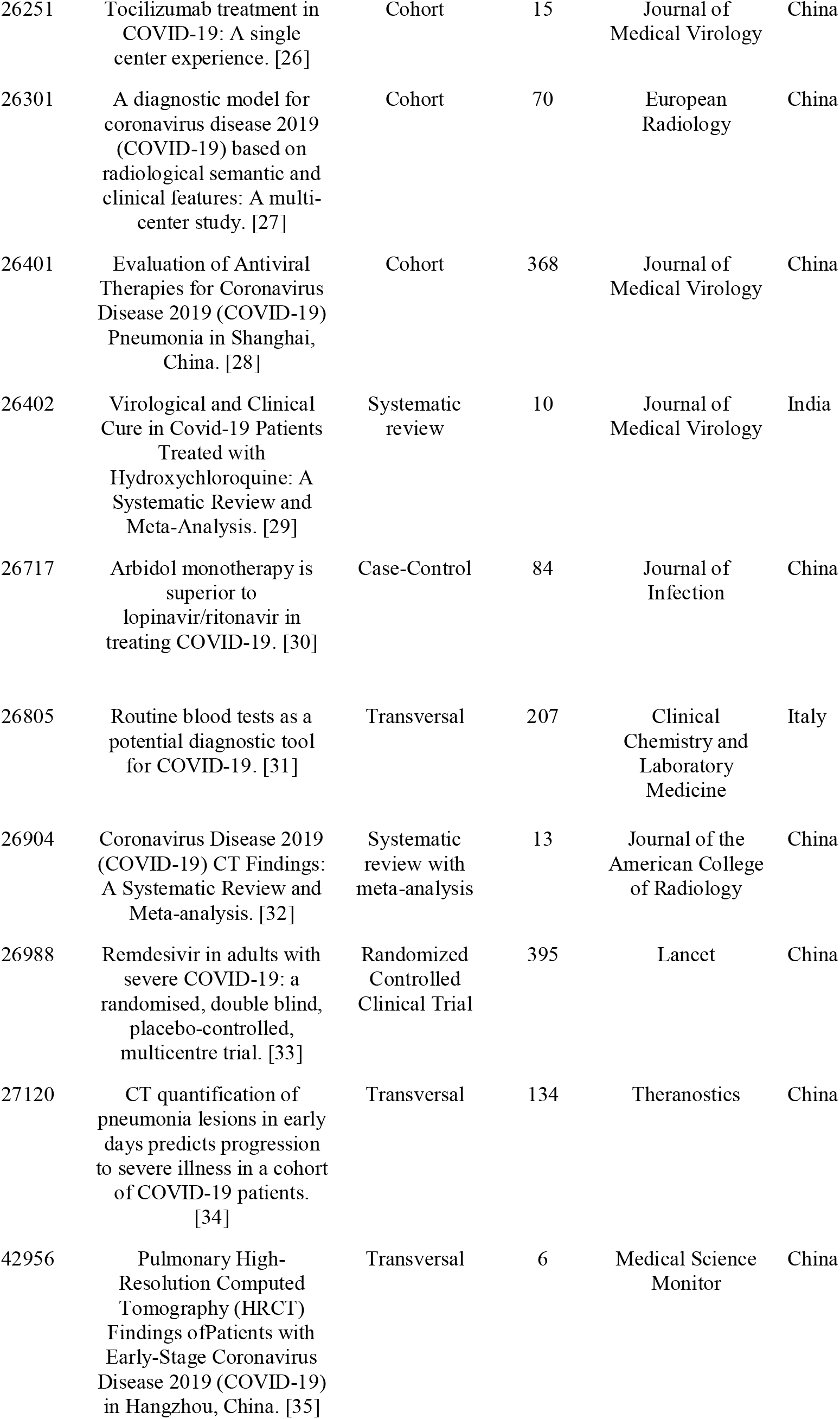
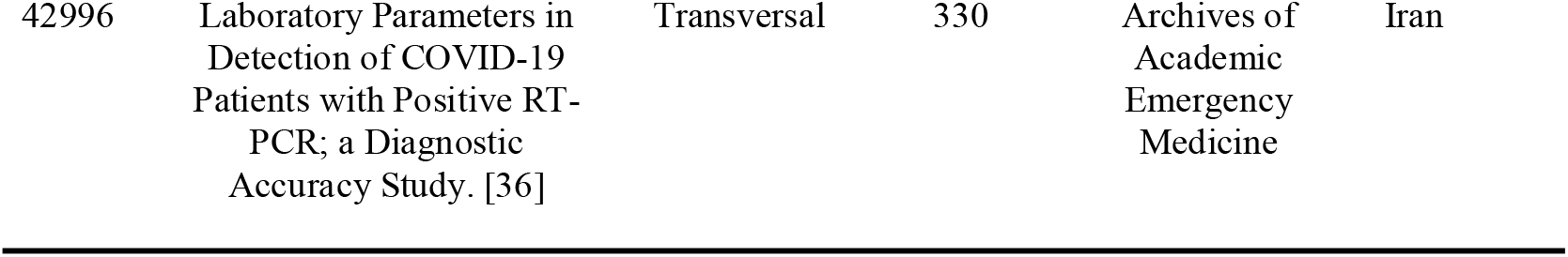
Characterization of the studies included in the revision, 2020 (n=30).

| ID | Title | Drawing | Sample | Journal | Country |
| --- | --- | --- | --- | --- | --- |
| 1830 | Diagnosis of the Coronavirus disease (COVID-19): rRT-PCR or CT? [7] | Case-Control | 36 | European Journal of Radiology | China |
| 1897 | Clinical efficacy of lopinavir/ritonavir in the treatment of Coronavirus disease 2019. [8] | Randomized Controlled Clinical Trial | 47 | European review for medical and pharmacological sciences | China |
| 1980 | Positive rate of RT-PCR detection of SARS-CoV-2 infection in 4880 cases from one hospital in Wuhan, China, from Jan to Feb 2020 [9] | Transversal | 4880 | Clinica Chimica Acta | China |
| 2045 | CT imaging features and image evolution characteristics of coronavirus disease 2019. [10] | Transversal | 57 | Journal of Central South University. Medical | China |
| 2099 | A Trial of Lopinavir-Ritonavir in Adults Hospitalized with Severe Covid-19. [11] | Randomized Controlled Clinical Trial | 199 | The New England Journal of medicine | China |
| 16332 | Association between chest CT features and clinical course of Coronavirus Disease 2019. [12] | Transversal | 195 | Respiratory Medicine | China |
| 16386 | Imaging and clinical features of patients with 2019 novel coronavirus SARS-CoV-2. [13] | Transversal | 90 | European Journal of Nuclear Medicine and Molecular Imaging | China |
| 16399 | Diagnostic value and dynamic variance of serum antibody in coronavirus disease 2019. [14] | Transversal | 76 | International Journal of Infectious Diseases | China |
| 16445 | Systematic review of the efficacy and safety of antiretroviral drugs against SARS, MERS or COVID- | Systematic review | 11 | Journal of the International AIDS Society | Switzerland |
| 19: initial assessment. [15] |  |  |  |  |  |
| 16526 | Clinical evaluation of a rapid colloidal gold immunochromatography assay for SARS-Cov-2 IgM/IgG. [16] | Cohort | 150 | American Journal of Translational Research | China |
| 16533 | Arbidol combined with LPV/r versus LPV/r alone against Corona Virus Disease 2019: A retrospective cohort study. [17] | Cohort | 56 | Journal of Infection | China |
| 24982 | Clinical evaluation of an immunochromatographic IgM/IgG antibody assay and chest computed tomography for the diagnosis of COVID-19. [18] | Transversal | 112 | Journal of Clinical Virology | Japan |
| 25217 | Tocilizumab therapy reduced intensive care unit admissions and/or mortality in COVID-19 patients. [19] | Caso-control | 20 | Médecine et Maladies Infectieuses | France |
| 25243 | Tocilizumab for the Treatment of Severe COVID-19. [20] | Cohort | 25 | Journal of Medical Virology | Qatar |
| 25876 | Plasminogen improves lung lesions and hypoxemia in patients with COVID-19. [21] | Almost experimental | 13 | An International Journal of Medicine | China |
| 26112 | Hydroxychloroquine and azithromycin as a treatment of COVID-19: results of an open-label non-randomized clinical trial. [22] | Non-randomized clinical trial | 48 | International Journal of Antimicrobial Agents | France |
| 26115 | Artificial Intelligence Distinguishes COVID-19 from Community Acquired Pneumonia on Chest CT. [23] | Transversal | 3322 | Radiology | China |
| 26171 | Correlation of Chest CT and RT-PCR Testing in Coronavirus Disease 2019 (COVID-19) in China: A Report of 1014 Cases. [24] | Cohort | 1014 | Radiology | China |
| 26211 | Corticosteroid treatment of patients with coronavirus disease 2019 (COVID-19). [25] | Transversal | 31 | The Medical Journal of Australia | China |
| 26251 | Tocilizumab treatment in COVID-19: A single center experience. [26] | Cohort | 15 | Journal of Medical Virology | China |
| 26301 | A diagnostic model for coronavirus disease 2019 (COVID-19) based on radiological semantic and clinical features: A multi-center study. [27] | Cohort | 70 | European Radiology | China |
| 26401 | Evaluation of Antiviral Therapies for Coronavirus Disease 2019 (COVID-19) Pneumonia in Shanghai, China. [28] | Cohort | 368 | Journal of Medical Virology | China |
| 26402 | Virological and Clinical Cure in Covid-19 Patients Treated with Hydroxychloroquine: A Systematic Review and Meta-Analysis. [29] | Systematic review | 10 | Journal of Medical Virology | India |
| 26717 | Arbidol monotherapy is superior to lopinavir/ritonavir in treating COVID-19. [30] | Case-Control | 84 | Journal of Infection | China |
| 26805 | Routine blood tests as a potential diagnostic tool for COVID-19. [31] | Transversal | 207 | Clinical Chemistry and Laboratory Medicine | Italy |
| 26904 | Coronavirus Disease 2019 (COVID-19) CT Findings: A Systematic Review and Meta-analysis. [32] | Systematic review with meta-analysis | 13 | Journal of the American College of Radiology | China |
| 26988 | Remdesivir in adults with severe COVID-19: a randomised, double blind, placebo-controlled, multicentre trial. [33] | Randomized Controlled Clinical Trial | 395 | Lancet | China |
| 27120 | CT quantification of pneumonia lesions in early days predicts progression to severe illness in a cohort of COVID-19 patients. [34] | Transversal | 134 | Theranostics | China |
| 42956 | Pulmonary High-Resolution Computed Tomography (HRCT) Findings of Patients with Early-Stage Coronavirus Disease 2019 (COVID-19) in Hangzhou, China. [35] | Transversal | 6 | Medical Science Monitor | China |
| 42996 | Laboratory Parameters in Detection of COVID-19 Patients with Positive RT-PCR; a Diagnostic Accuracy Study. [36] | Transversal | 330 | Archives of Academic Emergency Medicine | Iran |

### 2.3 Study selection

This review followed the guidelines of the Preferred Reporting Items for Systematic reviews and Meta-Analyzes extension for Scoping Reviews (PRISMA-ScR), according to the stages of identification, screening, eligibility, and inclusion of studies [6]. The stage of identification of the studies was carried out by three researchers together (S-AR, C-AB, and BM-C), and resulted in 6,060 articles, of which 190 were duplicated. The identified studies were organized in the Software State of the Art through Systematic Review (StArt), which contributed to the selection, screening, and extraction of data, according to the inclusion and exclusion criteria.

The inclusion and exclusion criteria were widely discussed among the researchers. Results of primary and secondary evidence, from the adult population over 18 years old, who presented methods of diagnosing COVID-19 and/or treating COVID-19 were included. The studies involving partial results on COVID-19, technical notes, preliminary reports, editorials, single case studies, approaches with a primary focus on pathophysiology, microbiology, or biochemistry were excluded.

The articles were screened based on two stages. In the first stage, an independent review was carried out by six pairs of researchers to reduce the potential bias (S-AR and AL; C-AB and C-CFR; DC and BM-C; M-EC and S -FRF; C-LGA and S-HS; M-AT and M-MF). A form for selection, sorting, and data extraction was developed in the StArt software and pre-tested by the researchers. Each researcher individually evaluated 980 titles and articles abstracts, approximately, regarding the inclusion or exclusion criteria, with subsequent issuance of the “accepted” or “rejected” opinion. Those articles that had no abstract available, their full text was accessed for screening. In cases where there was disagreement between the researchers of the pair, the article was referred to a third reviewer.

A total of 711 articles were eligible to read the full text. Kappa reliability, interobserver agreement was used. In the second stage of the screening, eleven independent researchers (S-AR; AL; C-AB; C-CFR; DC; M-EC; S-FRF; C-LGA; S-HS; M-AT; M-MF) performed the complete reading of the eligible articles. To reduce possible biases, five researchers (S-AR; A-L; C-AB; M-AT; C-CFR), together, evaluated the articles considered eligible. Everyone came together during this process to resolve the uncertainties related to the selection of the study. A total of 30 articles were finally included.

### 2.4 Charting the data

The form developed in the StArt software helped to extract data, such as country and year of publication, study design (descriptive, cross-sectional, case-control, cohort, ecological, randomized, quasi-experimental trial, systematic review, integrative review, or scoping review), confirmation of COVID-19 by laboratory tests (PCR and/or rapid test) and/or diagnostic imaging (X-ray, ultrasound, tomography and/or resonance), type of treatment of COVID-19 (antimalarial, anti-inflammatory, convalescent plasma, anticoagulant, antibiotic, corticoid, antiretroviral and/or others), drug name, dosage, efficacy (cure, death, clinical improvement, treatment change or ineffectiveness), results and outcomes, conclusions and/or recommendations.

### 2.5 Collating, summarizing and reporting the results

The extracted qualitative data were synthesized and organized into three categories, characterization of the included studies, diagnoses used for the detection of COVID-19, and associated treatments. Quantitative data regarding the geographic location of the literature and the type of approach, whether COVID-19 diagnosis and treatment or both, were georeferenced using ArcGIS Software version 10.6. The other data were analyzed using descriptive statistics, with the support of SPSS software version 20.0.

## 3. Results

According to the PRISMA statement, we initially selected 6.060 published articles. Among them, 190 articles were duplicated and finally excluded, resulting in 5.870 papers for the initial screening. After a careful screening and based on the scopus of the research, a total of 711 papers were included for full reading. Following the eligibility criteria, 30 papers were then adopted for the analysis (Figure 1). This study is presented in three categories as follows: characterization of the included studies, diagnoses used for the detection of COVID-19 and associated treatments.

**Figure 1.**
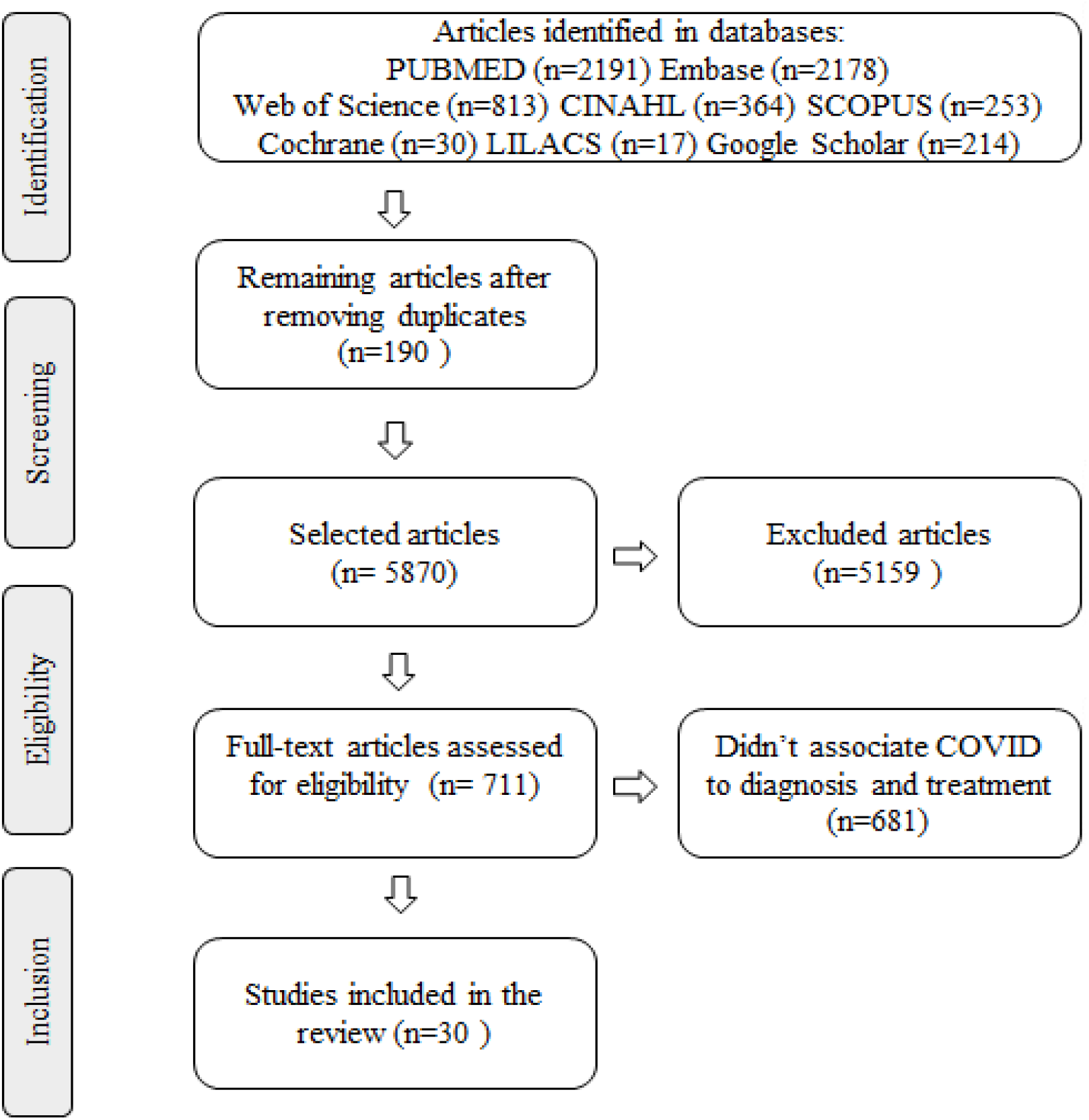
PRISMA flow diagram of assessment procedure and results: identification, screening, eligibility, inclusion and reasons of rejection.

### 3.1 Characterization of the included studies

After analyzing the spatial distribution of publications associated with COVID-19 in the world, it was noted that, until the time of data collection, there were few countries that had developed studies on the treatment and diagnosis of COVID-19. In Figure 2 we can verify that, the stronger the color degradation, the greater the publications and, in those countries in which they were not painted, there was no study available in the databases. The Figure 3 shows the publications related to diagnosis (n= 16; 53.33%) and treatments (n = 14; 46.67%). Therefore, China was the country that developed the most studies focusing in both diagnosis and treatment (n = 22; 73.33%) followed by France (n = 2; 6.67%); the Japan, Switzerland, Italy, Qatar, and India had one study published.

**Figure 2.**
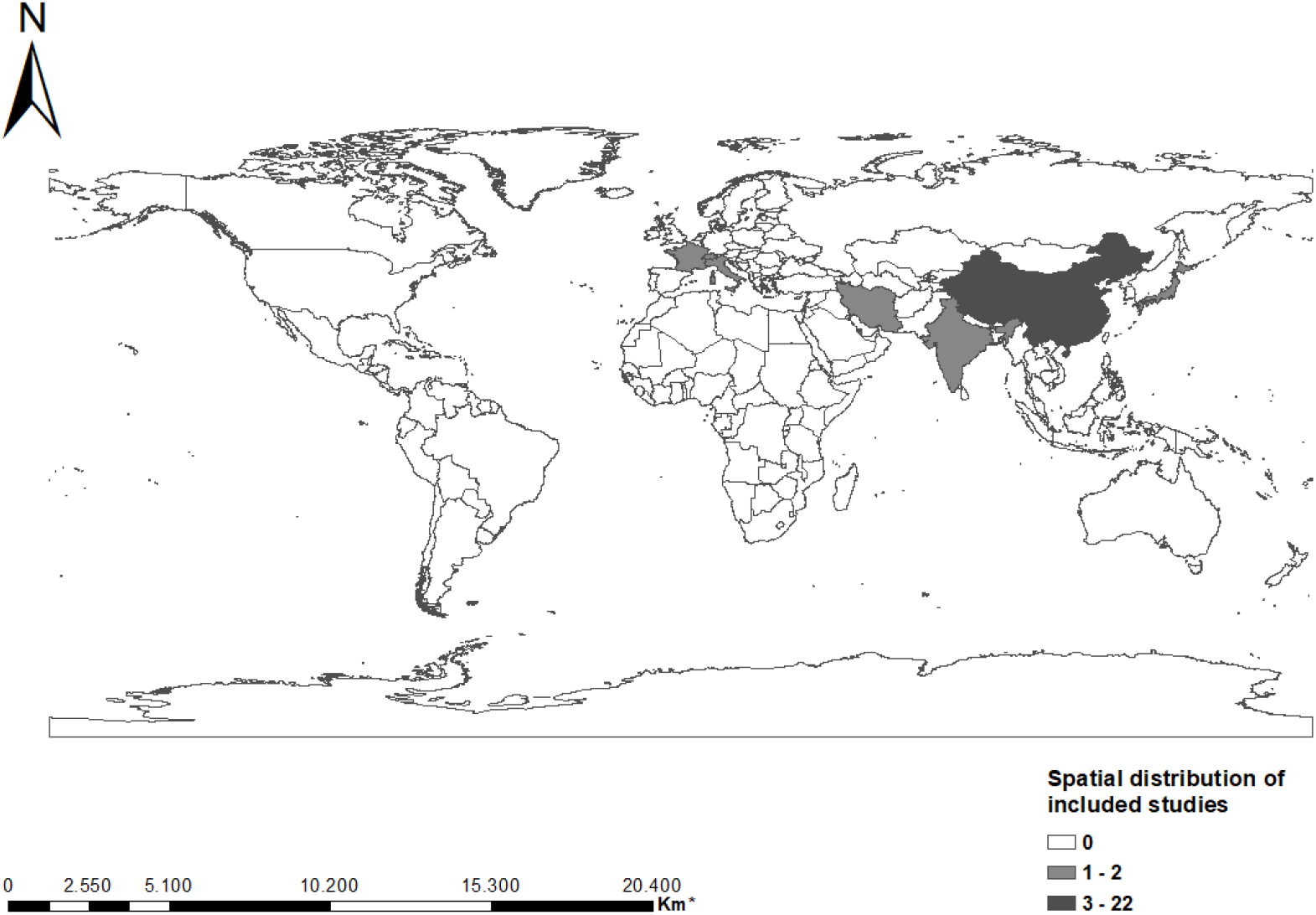
Spatial distribution of COVID-19 publications worldwide. * Graphical line segment scale indicates the measurement of distances on the map.

**Figure 3.**
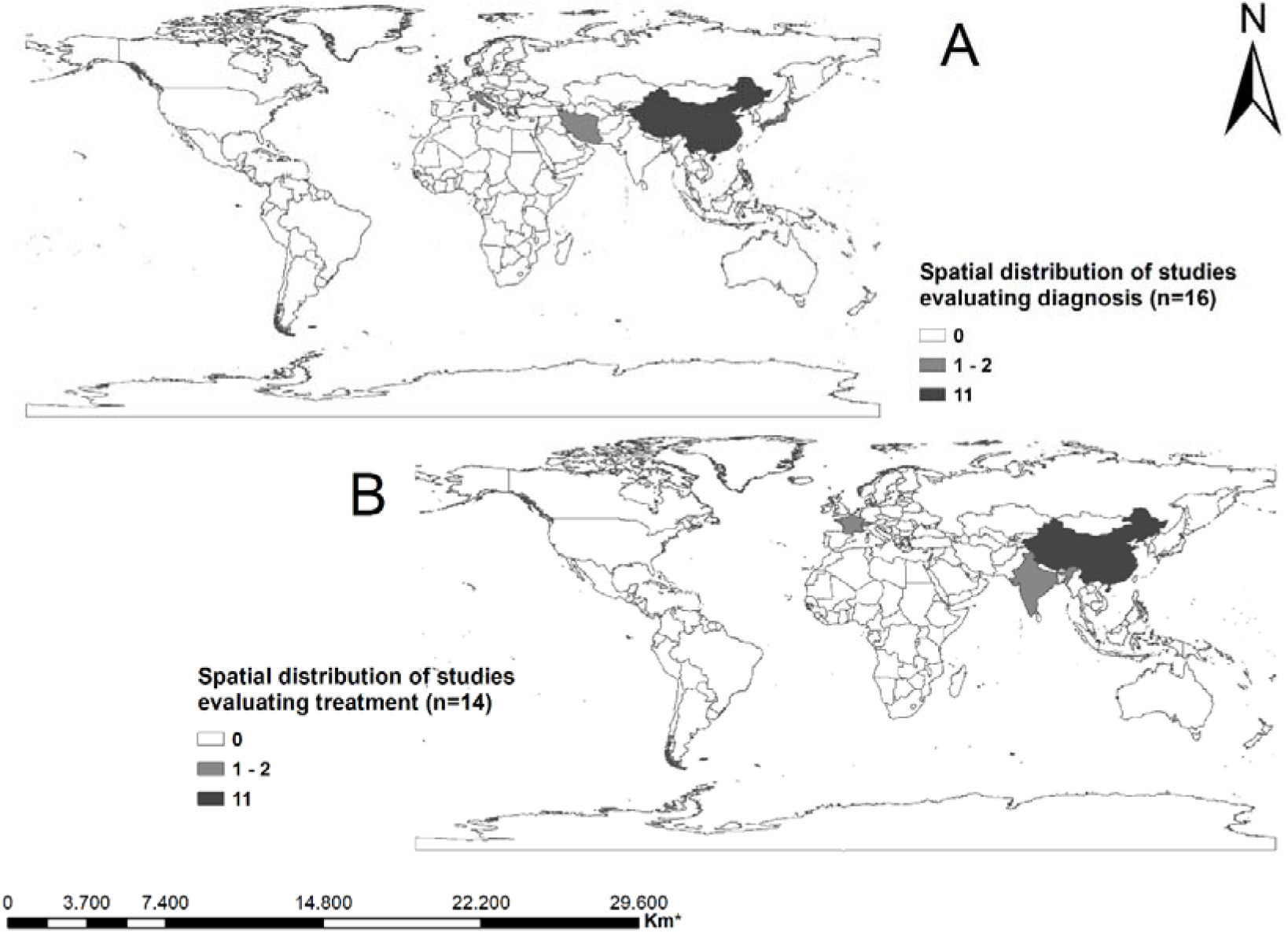
Spatial distribution of publications regarding both treatment and diagnosis of the COVID-19 in the world. * Graphical line segment scale indicates the measurement of distances on the map.

### 3.2 General characteristics of the included articles published in journals

All articles were peer-reviewed and published in 2020 (from january to late may). Of note, four articles (13.3%) were published in the Journal of Medical Virology, two articles (6.6%) in the Journal of infection and in Radiology, respectively; the other articles were included in different journals. A number of important and specific types of studies were included in this review, and the most prevalent was the Cross-sectional (n = 12, 40.0%), followed by Cohort (n= 7, 23.0%), Systematic Review (n= 3, 10, 0%) and Randomized Controlled Clinical Trial and Case-Control, both with totaling n = 3 (10.0%). The most used data base included Pubmed, Embase, and Web of Science (Figure 4 A) and based on data extraction we only accepted 4% of the total articles (Figure 4 B). The Figure 4 C depicts the arrangements of authors included in the scoping review. Using the number of appearance, we showed main location of publications (Figure 4 D); the amount of representative words were COVID-19, patients, and disease (Figure 4 E), and main descriptors were human, adult, article and female pandemic (Figure 4 F).

**Figure 4.**
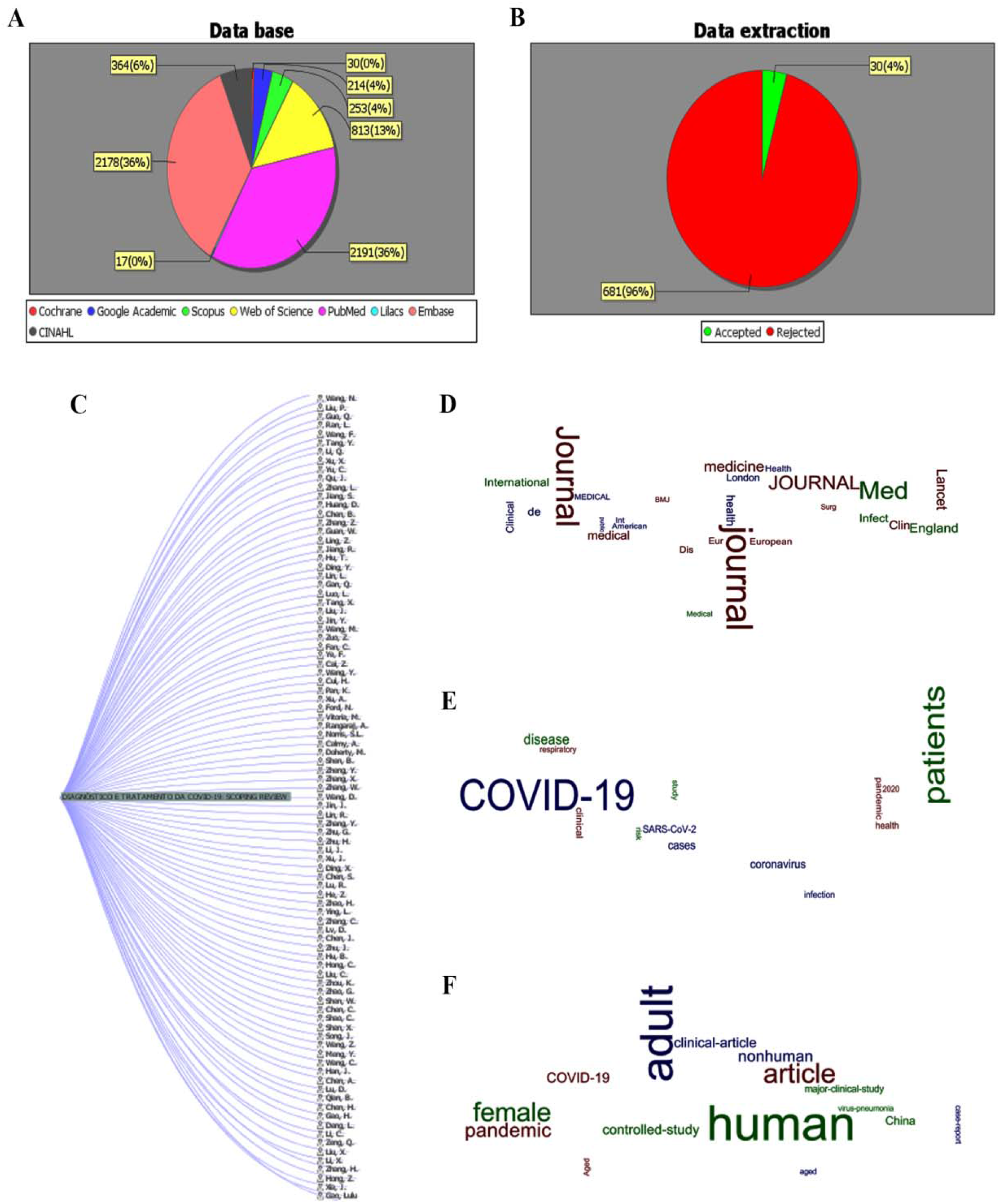
General features of the articles used in the review. A) Representative database in number and percentages; B) Data extraction of the 711 studies based on the second phase analysis; C) List of the authors that published articles involving COVID-19 diagnosis and/or treatments; D) Main locations of publications; E) Number of words appearing in the review; F) Main descriptors. Graphics provided by the StArt software.

Regarding the sampling of the included studies, the literature reviews included 10 to 13 articles, with an average of 11.33. Most notably, studies with human beings presented samples ranging from six to 4,880, with an average of 443.70 (Table 1).

### 3.3 Diagnostics used to detect SARS-CoV-2 infection

From these 30 articles, the majority of articles addressed the main subject “The diagnosis”. Among them, 58.8% evaluated the sensitivity of the diagnosis through computed tomography, and 41.2% evaluated the diagnosis via the laboratory test routine. Of these 30 articles included in this study, 16 articles addressed diagnosis. Of these, 58.8% (n=8) assessed the sensitivity of the diagnosis using computed tomography and 41.2% (n=7) assessed the diagnosis through routine laboratory tests (Table 2).

**Table 2.** Diagnostics associated with COVID-19. (N = 16)

| ID | Title | Objective | Conclusion |
| --- | --- | --- | --- |
| 1830 | Diagnosis of the Coronavirus disease (COVID-19): rRT-PCR or CT? [7] | To evaluate the diagnostic value of computed tomography (CT) and real-time reverse transcriptase-polymerase (rRT-PCR) chain reaction for COVID-19 pneumonia | RRT-PCR can produce initial false-negative results. It is suggested that patients with typical computed tomography findings, but with negative rRT-PCR results should be isolated, and rRT-PCR should be repeated to avoid diagnostic errors |
| 1980 | Positive rate of RT-PCR detection of SARS-CoV-2 infection in 4880 cases from one hospital in Wuhan, China, from Jan to Feb 2020. [9] | Retrospective analysis of Viral Nucleic Acid (NAT) tests of 4,880 cases from January 22 to February 14, 2020, at Renmin Hospital, Wuhan University | The viral nucleic acid (NAT) test played an important role in identifying SARS-CoV-2 infection |
| 2045 | CT imaging features and image evolution characteristics of coronavirus disease 2019. [10] | To analyze the imaging characteristics of COVID-19 in different periods, and resume the characteristics with its development | The initial manifestations of COVID-19 CT scan are mainly foci of density in ground glass, distributed in the subpleural region, some are distributed close to the bundle of bronchial blood vessels and in the central area of the lobe |
| 16332 | Association between chest CT features and clinical course of Coronavirus Disease 2019. [12] | To illustrate the radiographic characteristics of Coronavirus Disease 2019 and the correlation with clinical evolution | There are several specific image changes, along with disease progression, that can be useful in early recognition and differential diagnosis of Coronavirus Disease 2019 |
| 16386 | Imaging and clinical features of patients with 2019 novel coronavirus SARS-CoV-2. [13] | To report the clinical and imaging characteristics of patients infected with SARS-CoV-2 in Guangzhou, China | The CT scan of the chest plays an important role in the initial diagnosis of COVID-19 pneumonia. Multiple ground-glass opacities in the bilateral lobe with peripheral distribution are typical of computed tomography in case of COVID-19 pneumonia |
| 16399 | Diagnostic value and dynamic variance of serum antibody in coronavirus disease 2019. [14] | To investigate the diagnostic value of serological testicles and dynamic variance of serum antibody in COVID-19 | The serological test is effective for the diagnosis of SARS-CoV-2 infection. The positive rate and variance of the IgG titer are higher compared to IgM in COVID-19 |
| 16526 | Clinical evaluation of a rapid colloidal gold immunochromatography assay for SARS-Cov-2 IgM/IgG. [16] | To assess the potential to be used in screening patients with COVID-19 | The colloidal gold immunochromatography assay for the SARS-Cov-2 specific IgM/IgG antibody has a sensitivity of 71.1% and specificity of 96.2% in this population, showing potential as a rapid diagnostic test |
| 24982 | Clinical evaluation of an immunochromatographic IgM/IgG antibody assay and chest computed tomography for the diagnosis of COVID-19. [17] | To evaluate the clinical performance of an IgM / IgG immunochromatographic (IC) assay for severe acute respiratory syndrome coronavirus 2 (SARS-CoV2) and chest computed tomography (CT) for the diagnosis of 2019 coronavirus disease (COVID-19) | IC assay had low sensitivity during the early stage of infection and therefore the assay alone is not recommended for the initial diagnostic test for COVID-19. If RT-qPCR is not available, the combination of chest CT and IC assay can be useful in diagnosing COVID-19 |
| 26115 | Artificial Intelligence Distinguishes COVID-19 from Community Acquired Pneumonia on Chest CT. [22] | To develop a fully automatic framework to detect COVID-19 using CT tomography its performance. | A deep learning model can accurately detect COVID-19 and differentiate it from community-acquired pneumonia and other lung diseases. |
| 26171 | Correlation of Chest CT and RT-PCR Testing in Coronavirus Disease 2019 (COVID-19) in China: A Report of 1014 Cases. [24] | To investigate the diagnostic value and consistency of chest CT compared to the RT-PCR assay in COVID-19 | A highly sensitive CT for the diagnosis of COVID-19. Chest CT can be considered the main tool for the current detection of COVID-19 in epidemic areas |
| 26301 | A diagnostic model for coronavirus disease 2019 (COVID-19) based on radiological semantic and clinical features: a multi-center study. [27] | To identify differences in CT images and clinical manifestations between patients with pneumonia with and without COVID-19, and to develop and validate a diagnostic model for COVID-19 based only on clinical characteristics and radiological semantics | Based only on CT images and clinical manifestations, patients with pneumonia with and without COVID-19 can be distinguished. A model composed of radiological and clinical semantic characteristics has excellent performance for the diagnosis of COVID-19 |
| 26805 | Routine blood tests as a potential diagnostic tool for COVID-19. [31] | To evidence statistically relevant differences that may be useful in identifying positive and negative COVID-19 patients in plasma leukocyte (WBCs), platelets, C-reactive protein (CRP), aspartate aminotransferase (AST), alanine aminotransferase (ALT), γ-glutamyl | A combination of cut-off points for hematological parameters can help identify false positive/negative rRT-PCR testicles. A blood test analysis can be used as an alternative to rRT-PCR to identify patients positive for COVID-19 in countries that serve with a large shortage of reagents for rRT-PCR |
|  |  | transpeptidase (GGT), alkaline phosphatase (ALP) and lactate dehydrogenase (LDH) | and/or a specialized laboratory |
| 26904 | Coronavirus Disease 2019 (COVID-19) CT Findings: A Systematic Review and Meta-analysis. [32] | To provide a summary of the evidence on the detection of COVID-19 by chest CT and as expected CT image manifestations | The detection of chest computed tomography in COVID-19 is very high among those subject to high-risk symptoms, mainly thin section chest CT. The most common features of CT in patients affected by COVID-19 included ground-glass opacities and encompassing the bilateral lungs in a peripheral distribution |
| 27120 | CT quantification of pneumonia lesions in early days predicts progression to severe illness in a cohort of COVID-19 patients. [33] | To quantify pneumonia lesions by computed CT in the first days to predict progression to severe disease in a cohort of patients with COVID-19 | The CT quantification of pneumonia lesions can predict early and non-invasive progression to severe disease, providing a promising prognostic indicator for the clinical treatment of COVID-19 |
| 42956 | Pulmonary High-Resolution Computed Tomography (HRCT) Findings of Patients with Early-Stage Coronavirus Disease 2019 (COVID-19) in Hangzhou, China. [35] | To investigate the imaging manifestations of early-stage coronavirus disease 2019 (COVID-19) and provide an imaging base for early detection of suspected cases and stratified intervention | This study revealed that a CT scan performed in the initial stage of the disease shows some missed diagnoses, so it is produced in short that it is used together with the other diagnostic means. |
| 42996 | Laboratory Parameters in Detection of COVID-19 Patients with Positive RT-PCR; a Diagnostic Accuracy Study. [36] | To evaluate the accuracy of laboratory parameters in predicting cases with positive RT-PCR for COVID-19 | Based on the results, ALT, PCR, NEU, LDH, and Urea have very good accuracy in predicting cases with COVID-19 positive RT-PCR |

#### 3.3.1 Computed tomography (CT) as diagnostic imaging test

CT was considered as the sensitivity factor for the detection of COVID-19, resulting in percentages above 90.0% (ID: 1830, 26171 and 26115). Based on CT and clinical manifestations, it is possible to distinguish patients with pneumonia presenting COVID-19 or not (ID: 26301). There was a predictive capacity regarding the tomography resources. After analyzing the 1^st^ and 4^th^ days after admission, it is easy to predict early and non-invasively the progression to severe disease, providing a promising prognostic indicator for the clinical management of COVID -19 (ID: 27120).

When analyzing the tomography images, most patients had peripheral lesion distribution (ID: 2045, 16332, 16386, 26301, 26904), as well as ground glass density foci (ID: 2045, 16332, 16386, 26301, 26904, 42956). There was also an association with advanced age, resulting in higher case number and in greater complexity of the pathology (ID: 16332, 26115 and 27120).

#### 3.3.2 Diagnostics by laboratory tests

We observed that the serological samples showed sensitivities to the IgM and IgG antibodies for the diagnosis of COVID-19 (ID: 16399 and 16526). In addition to an important sensitivity rate of 71.1% (ID: 16526) and 100% and 90.9%, the detection rate of positive IgG was higher than IgM (ID: 16399). Another study documented that a combination of CT and immunochromatographic assay could be complementary, since this combination presents a sensitivity rate of 82.4% in symptomatic cases; it should be emphasized that due to the low sensitivity (ID: 24982), the collection of isolated immunochromatography is not recommended for initial testing.

Three studies evaluated the RT-PCR (ID: 1980, 42996 and 26805). After analyzing the detection of SARS-CoV-2 by RT-PCR of the viral nucleic acid in a population of 4,880 patients, the positive rate was 38% (n = 1875) and the samples of nasal and pharyngeal swabs (n = 4818) had a positivity rate of 38.25% and 49.12% in sputum, thus showing an important role in the identification of SARS-CoV-2 infection (ID: 1980).

When assessing the accuracy of laboratory parameters in predicting cases with RT-PCR, a total of 200 cases (n = 70; 35%) were positive; they revealed significantly higher number of neutrophil (NEU) (p = 0.0001) and changes in the serum levels of C-reactive protein (PCR) (p = 0.04), lactate dehydrogenase (LDH) (p = 0.0001), aspartate aminotransferase (AST) (p = 0.001), alanine aminotransferase (ALT) (p = 0, 0001) and urea (p = 0.001). They also had lower white blood cell (leukocyte) count (p = 0.0001) and serum albumin level (p = 0.0001). Of note, LDH, CRP, ALT and NEU may be used to predict the result of the COVID-19 test, thus providing us a more precise detection method (ID: 42996).

An interesting study sought to compare two groups of patients with positive rRT-PCR results (positive group) and 102 patients with negative rRT-PCR results (negative group), and it was indicated a strong association between COVID-19-positive patients with a low blood leukocytes (neutrophils, eosinophils, and basophils), lymphocytes and monocytes in the positive rRT-PCR group. Finally, elevation in activities of pyridoxal phosphate-dependent enzymes, AST and ALT were also observed in the positive group (ID: 26805).

### 3.4 Treatments used to fight SARS-CoV-2 infection

We registered and included 14 articles regarding the COVID-19 treatment as the main subject. The most common treatments included the Lopinavir/Ritonavir (LPV/r), Arbidol, Hydroxychloroquine, Azithromycin, Corticosteroids, and Plasminogen as therapeutic forms (Table 3).

**Table 3.**
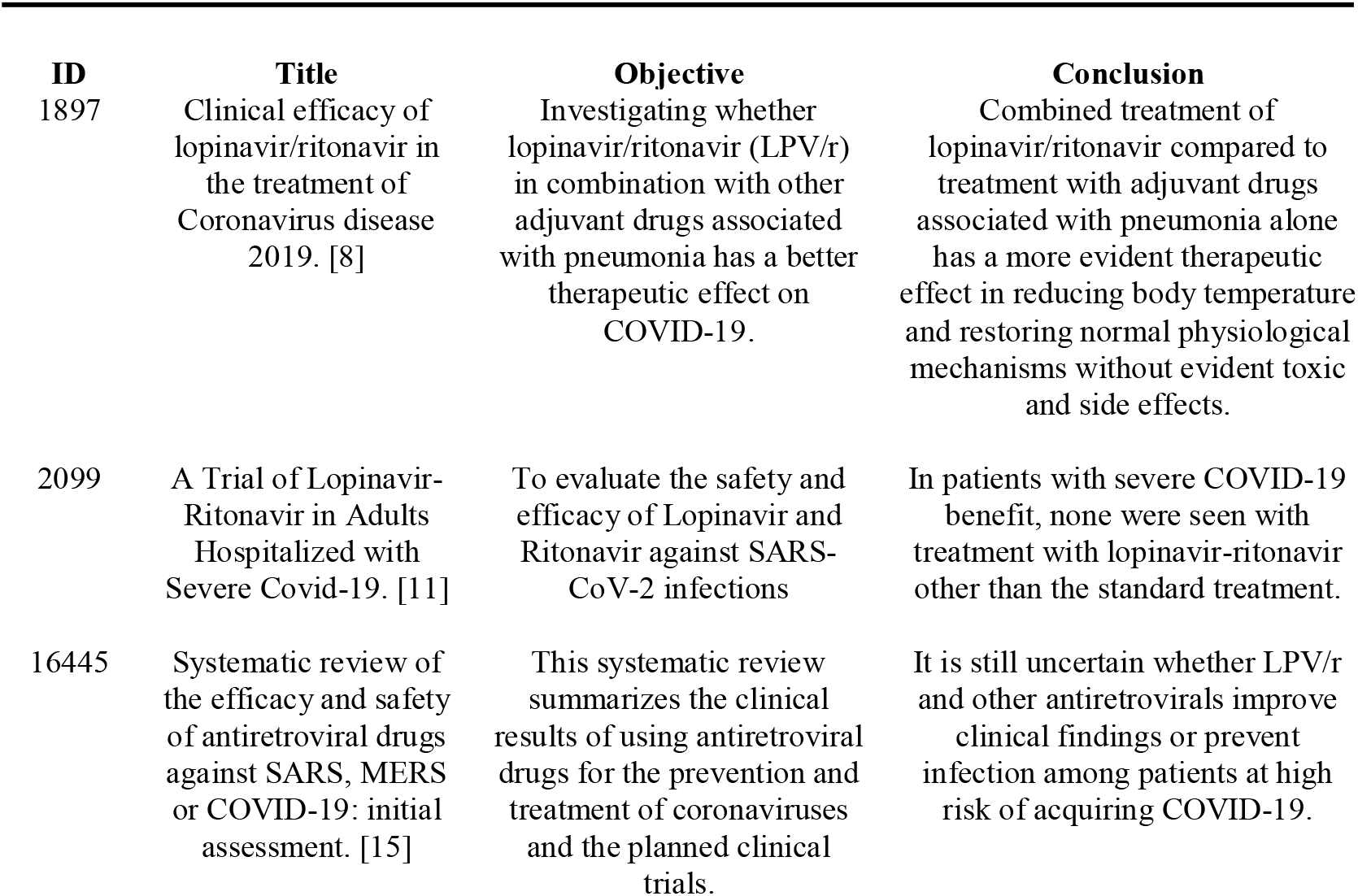
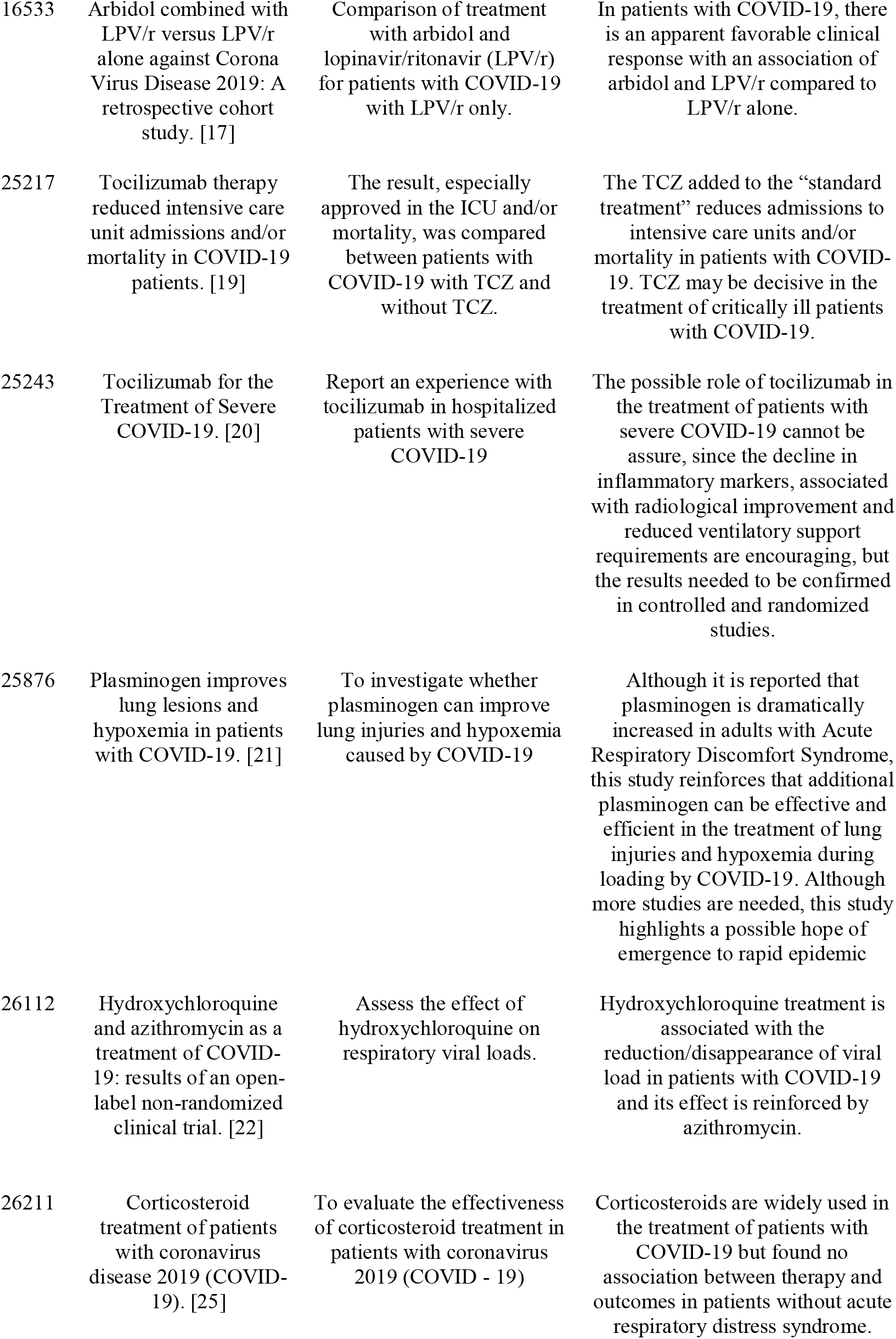
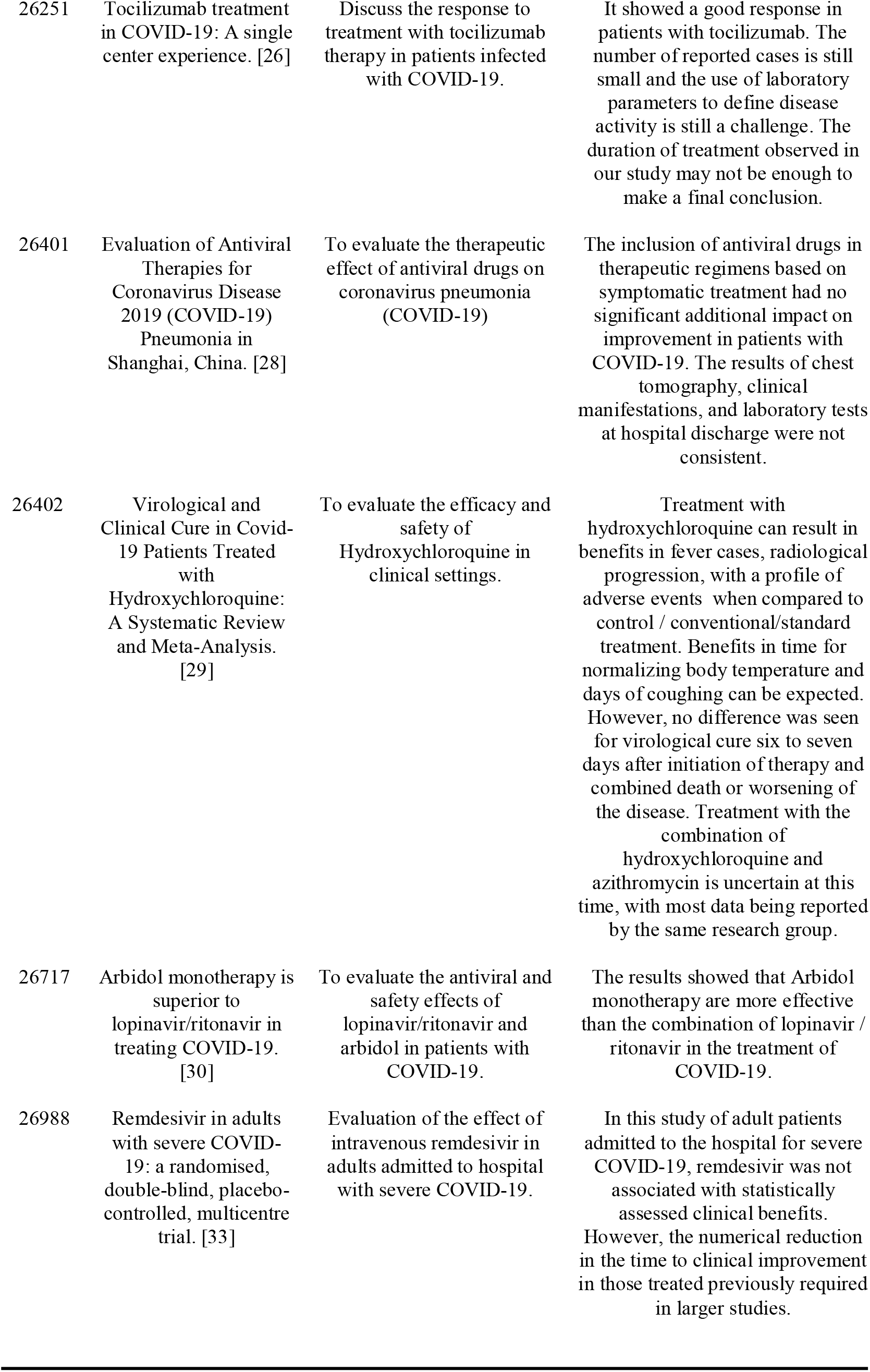
Main treatments associated with COVID-19. (n = 14)

#### 3.4.1 Lopinavir / Ritonavir (LPV/r) and Arbidol

The studies that reported the effect of using Lopinavir / Ritonavir (LPV/r) pointed to several controversies and uncertainties about the effectiveness of the combination added to the standard treatment in people with Covid-19 (ID: 2099, 16445, 1897 and 26401). Regarding the use of adjuvant drugs associated with pneumonia alone, the treatment with LPV/r showed a more evident therapeutic effect in reducing symptoms such as body temperature and in restoring normal physiological mechanisms. These therapeutic regimens showed no evident toxic/side effects and presented improvements in relation to laboratory results; they acted by reducing the abnormal proportion of white blood cells, lymphocytes, and C-reactive protein in COVID-19-infected patients (ID: 1897). Considering the mortality rate in 28 days, both standard and combined treatment (LPV / r) were similar (19.2% vs. 25.0%; 95% CI). In addition, some adverse effects such as nausea, vomiting and diarrhea were more common in patients treated with LPV / r (ID: 2099).

A systematic review identified that administration of LPV/r compared to standard treatment did not show a significant difference in the time of clinical improvement (ID: 26401 and 16445), although the treated group showed the shorter time. Notably, the LPV/r-treated group displayed a lower mortality (ID: 16445) than that of standard treatment. The review highlights two studies describing a possible protective effect of LPV/r as profilaxis after viral exposure. In a cohort study, no differences were found between LPV/r-treated group and the standard treatment for the controlo of pneumonia and clinical results (ID: 26401).

Other articles described the effect of Arbidol and LPV/r (ID: 16533, 26717), one of which evaluated the association of Arbidol and LPV/r compared to those treated with LPV / r only (ID: 16533) while other evaluated the combination of LPV/r with Arbidol monotherapy (ID: 26717). Both articles showed better results for the groups that either used Arbidol, in combination with LPV/r or even used as monotherapy. When comparing the use of Arbidol + LPV/r with isolated LPV/r, it was observed that chest CT images improved in 69.0% of patients who used the combination Arbidol + LPV. Furthermore, the detection rate of SARS-CoV-2 in nasopharyngeal samples after seven days of treatment was reduced in the group using the triple combination.

By investigating these compounds affecting the viral loading, the authors demonstrated a 100.0% reduction in the detection of viral load after 14 days of admission in patients treated with Arbidol (n = 16); otherwise, in the group treated with LPV/r (n = 34), the viral load was found in 44.1% of patients (ID: 26717). On the other hand, a cohort that evaluated the use of monotherapy with Arbidol found no clinical efficacy when compared with patients treated only for their symptoms (ID: 26401). This same study observed no improvements in pulmonary images or in the average length of hospital stay after combining with other antivirals (Arbidol + Ritonavir or Interferon + LPV/r or Interferon + Darunavir).

#### 3.4.2 Hydroxychloroquine and Azithromycin

Patients treated with Hydroxychloroquine (HCQ) (ID: 26402) demonstrated a reduction in radiological progression of the disease compared to conventional treatment (OR for radiological disease progression during treatment was 0.31; 95% CI, [0, 11-0.9]). In addition to these results, there were signs of benefits of HCQ over time for normalizing body temperature while reducing the number of days of dry coughing. Through comparison of patients treated with HCQ and the controls (treated with conventional treatment), no expressive differences in virological cure (OR, 2.37, 95% CI, 0.13-44.53), death or clinical worsening of the disease (OR, 1.37, 95% CI, 1, 37-21.97) and safety (OR, 2.19, 95% CI, 0.59-8.18) was observed. Coversely, a clinical trial (ID: 26112) demonstrated a significant reduction or elimination of viral load; after six days of treatment inclusion, 70% of the patients were cured virologically compared to the control group (12.5%, p = 0.001).

Regarding the adverse effects of HCQ, a total of seven events were identified in the systematic review and included nausea, diarrhea, abnormal liver function, skin rash and headache; however, when these results were combined and analyzed, no significant difference was observed between the two arms (OR, 2.19; 95% CI, [0.59-8.18]) (ID: 26402). Other relevant studies investigated the combinatory effect of HCQ with Azithromycin (ID: 26402 and 26112), and according to the systematic review, five studies reported both the safety and the effectiveness of this combination (ID: 26402). More importantly, the clinical trial showed that after five days of treatment with HCQ + Azithromycin, 100.0% of the patients presented with virological cure compared with 50.0% in those treated with HQC alone and 18.8% in the control group (p = 0.002) (ID: 26112).

#### 3.4.3 Tocilizumab e Remdesivir

Three articles reported the use of Tocilizumab in people with COVID-19 (ID: 25217; 25243 and 26251) and concluded that treatment with Tocilizumab significantly reduced the proportion of patients on invasive mechanical ventilation (ID: 25217 and 25243) and the number of death outcome and admissions to the Intensive Care Unit (ID: 25217). In addition, there was a significant reduction in C-reactive protein levels and benefits in decreasing inflammatory activity (ID: 26251). The adverse events associated with Tocilizumab were anemia, increased alanine aminotransferase and prolonged QT interval (ID: 25243).

Regarding the use of Remdesivir, no time difference was observed until clinical improvement (risk ratio 1 23 [95% CI 0 87-1 75]). In addition, the adverse events were reported in 66% of treated patients, being the most described as constipation, hypoalbuminemia, hypokalemia, anemia, thrombocytopenia, and increased total bilirubin. Although not significant, the time to clinical improvement was faster for patients using Remdesivir (ID: 26988)

#### 3.4.4 Corticosteroids and Plasminogen

By evaluating the efficacy of corticosteroids (Methylprednisolone) used in the treatment of COVID-19, it was not possible to verify an association between the treatment and the time of virus elimination, length of hospitalization or duration of symptoms. In fact, patients treated with corticosteroids had more clinical symptoms, a higher rate of inflammation and several abnormalities on chest tomography (ID: 26211). The inhalation of freeze-dried plasminogen was also evaluated in patients with COVID-19 and the main results for clinically moderate patients included improvements in lung injury conditions and reduction in the heart frequency; patients with more severe conditions tends to show better oxygen saturation (ID: 25876). This may be effective and efficient in the treatment of lung injuries and hypoxemia during infections by SARS-CoV-2.

## 4. Discussion

We conducted a scope review of COVID-19, thoroughly researching databases and other sources based on the geographic distribution of publications until May 2020, and, China was the country with the largest number of publications related to COVID-19. Our scoping review consistently evaluated important studies that emerged during the first semester in an attempt to explore what is expected in terms of diagnosis and treatment. Facing this challenge, we incorporated several study types and research areas, with special attention to the randomised controlled trials and cohort studies, which were non-existent before early February.

To date, the main diagnostic methods described included CT imaging and laboratory testing. It is assumed that imaging methods are effective to differentiate the degree and the involvement of the respiratory organs by the virus following patients stratification with various ages. In fact, the great majority of studies performed in hospitals and specialized clinics were dedicated to explore CT findings. Likewise, the laboratory tests using serum or nasopharyngeal samples of patients are widely used to detect produced antibodies, enzymatic activities or the viral counterparts. It seems true that the combination of CT images with RT-PCR provide a more realistic framework to detect the SARS-CoV-2 infection with disease aggravation. Other important parameters that should be taken into consideration after viral infection confirmation are the number of inflammatory and immune cells in association with the levels of PCR, LDH, AST, ALT, albumin, and urea.

The research topics found in the articles/studies had both similarities and differences. Basic research was mostly settled on correlating the COVID-19 diagnosis with altered parameters and molecular signatures in a number of tissues and systems; for the current scoping, this was often filtered out so that we only adopt research on healthy patients to avoid sampling bias.

It seems true that SARS-CoV-2 has also been detected in non-respiratory specimens, including blood, ocular fluids, stool, and semen; however, if these sites predispose to the transmission is still unclear [37–41]. Although the detection of SARS-CoV-2 RNA in blood has also been recently reported not all studies have tested it [42,43]. Our review did not retrieve studies on detection of short- and long-term risks of reinfection. Despite the recent data on individuals tested for reinfection exists and follows a distinct paradigm [44,45], it is important to employ a combination of strategies, i.e., images and blood dosages, to map these immunological differences.

This review may provide to other researchers identification of contradictory results and research gaps allowing future approaches to fill it. Notably, an integrative review matching the diagnostic methods with personal parameters throughout disease evolution would be of great value. The reviews and short reports generally provide generic information of the virus, transmission, and treatments. A recent scoping review (from 01 december to 06 February) described in detail the evidence for the development of clinical practice guidelines and public health policies [46]; in that time, the authors had few clinical research available. In this sense, our scoping review may add significant contribution to this study since it was centered in clinical practice advances and in medical management of hospitalized patients. On one hand, our scoping review has weakness regarding few studies and variability of methods and reported data. On the other hand, all studies have many strengths as they were well-conducted using large sample size and with high potential for reproducibility.

The effectiveness of the treatments for COVID-19 are still controversial and open for tireless debate. Based on our scoping review, different drugs were tested, ones with better outcomes than others. During the period we evaluated the studies, there were no therapeutic regimen with specific dosage or duration that can be applied to every patient; so far, a vaccine is not available. We now summarize and discuss the main results about the treatments we have surveyed.

Patients treated with the HIV combined therapy Lopinavir (PubChem CID: 92727) and Ritonavir (PubChem CID: 392622) had improved laboratory results, but the mortality rate did not differ between LPV/r and the standard protocol (ID: 26401 and 16445). Besides, the certainty of the evidence of randomized and observational studies were frequently low. Corroborating this issue, a randomized, controlled, open-label trial with 199 adult patients found no additional benefits with LPV/r treatment compared to standard supportive care alone, *i.e*., oxygen and vasopressor support, antibiotics, renal-replacement therapy and extracorporeal membrane oxygenation. Also, after 28 days of the combined treatment the viral RNA loads or duration of viral RNA detectability were similar between patients in the LPV/r and in the standard care group [11]. On the other hand, studies evaluating the combination of LPV/r with Arbidol (PubChem CID: 31411) or Arbidol monotherapy had distinct outcome (ID: 16533, 26717), making Arbidol, also known as Umifenovir, a promising repurposed candidate to treat COVID-19. In a small cohort with 67 patients admitted with abnormal chest CT findings, Arbidol treatment tended to increase the discharging rate and reduced mortality [47]. To confirm the efficacy of the treatment with Arbidol there is an ongoing randomized, open, multicenter clinical trial scheduled to publish its outcomes in December of the current year (NCT04260594). Anyway, the findings of our review lead us to believe that the combination of Lopinavir, Ritonavir and Arabidol may help in delaying the progression of lung injuries while decreasing the likelihood of respiratory and gastrointestinal transmission, and avoiding long-standing hospitalization as well. However, we do not rule out the need for new randomized, multicenter research with larger samples.

Although chloroquine (PubChem CID:3652) and its safer derivative, hydroxychloroquine (PubChem CID:3652) have already a well-documented history for coping malaria and inflammatory autoimmune disease, these drugs are under the spotlight of a heated debate about their safety, efficacy and cost benefit for the treatment of COVID-19. For the sake of contextualize hydroxychloroquine has a more potent *in vitro* activity, *i.e*., lower EC_50_, and is more effective than chloroquine for both prophylaxis and treatment [48]. The study by Sarma and colleagues demonstrated that HCQ improved the radiologic findings, shortened the periods of fever and dry coughing, however, it was not observed significantly difference in the viral clearance compared to the control group. A prospective randomized study of 30 patients in China showed that after seven days receiving HCQ plus standard cares, virologic clearance was similar to those who received the standard care alone [49]. On the other hand, Gautret et al. [50] demonstrated the efficiency of HCQ in clearing viral nasopharyngeal carriage and went farther showing that, within five days, the combination of HCQ with the antibiotic Azithromycin (PubChem CID:447043) negativized 6 of 6 patients infected with the SARS-CoV-2 *versus* 8 of 14 patients that received HCQ alone. Weighing up the pros and cons of the possible therapeutic regimens with chloroquine or HCQ, one may argue that these drugs are relatively well tolerated and no significant adverse effects have been reported at the doses and durations proposed for COVID-19 treatment [51]; on the flip side, it should not be disregard the serious possible adverse effects that, though rare, include QTc prolongation, hypoglycemia, neuropsychiatric effects, and retinopathy [52]. Furthermore, there were few statistically robust results confirming the regimen efficacy. Together, these evidence lead us to suggest that before initiating the treatment for COVID-19, it is mandatory an individualized, detailed, independent and, above all, consensual assessment to make the best possible choice.

When the therapy was conducted with Tocilizumab (PubChem SID: 135345962) the number of patients that were admitted to Intensive Care Unit and needed invasive mechanical ventilation was reduced (ID: 25217; 25243 and 26251). From those patients who were already in ICU, 76% improved or remained stable after Tocilizumab treatment. Overall, from 100 patients admitted in one study, 77 of them benefited from the treatment. However, 23 patients had their respiratory condition worsened and 20 of them died [53]. Contrasting this study, a cohort conducted by Campochiaro et al. [54] verified no clinical improvement or reduced mortality between tocilizumab and standard treatment patients. The use of Remdesivir (PubChem CID: 121304016) did not improve clinical outcomes and was associated with some adverse effects. Therefore, we reinforce the need for more literature about the definitive efficacy of the humanized monoclonal antibody that targets the IL-6 receptor, and about the ATP analog. More recently, the solidary big trial by WHO, which enrolled approximately 11,000 individuals in 400 hospitals around the world, revealed that hydroxychloroquine, ritonavir/lopinavir, and remdesivir did not increase patients survival nor lowered mortality or delayed the urgent need for artificial ventilation [55].

Although there is enough evidence that inflammatory status plays a determinant role in the clinic evolution of patients with COVID-19, a study (ID: 26211) analyzing the use of methylprednisolone (PubChem CID: 6741) showed no benefit, but only adverse effects. Conversely, 26 patients treated with low-dose of corticosteroid for a few days had faster recovery and improvement of lung symptoms [47]. According to Saghazadeh et al. [56], the use of corticosteroids may cause suppression of antiviral immune response, so the empiric use of this class of drug may be restricted until controlled clinical trials prove that this treatment modality ameliorates the inflammatory-related symptoms and reduces the COVID-19-related death. The inhalation of manipulated plasminogen was useful in treating lung lesions and hypoxemia, however this was demonstrated by only one study, and the heterogeneity of the patients (clinically moderate, severe or critical) reinforces the need for further trials.

## 5. Conclusion

The main evidence related to diagnostic methods is clear, and includes tomography and laboratory tests. However, we felt a lack of rigorous studies focused on novel and more reliable diagnostics methods. The medications for the treatment of COVID-19, although showing some reduction of the signs and symptoms related to this disease, the viral load, inflammatory activity and mortality, may cause adverse effects of mild, medium or severe intensity. More studies are encouraged to continuously review and update the literature on this subject to effectively uncover a feasible therapy to fight COVID-19 until the vaccine is released safely and affordable.

## Data Availability

Data and materials are fully available without restriction

## Financial disclosure

This research did not receive any specific grant from funding agencies.

## Availability of supporting data

Data and materials are fully available without restriction

## Conflicts of Interest

The authors declare no conflict of interest.

## Author Approval

All authors approved the final manuscript

## Author contributions

ARS, ECM: conception and design the study, analysis and interpretation of data. CBM, CFC, LRA, CD: analyzed the data, final review. FRFR, MFM, AM: participated in the acquisition and interpretation of data, and in the intellectual conception of the study. LGAC, HSS: design, critical analysis and draft the manuscript. The authors approved the final version of the manuscript.

## Notes

### Competing Interest Statement

The authors have declared no competing interest.

### Author Declarations

This work is a scoping review of the literature. Not applicable.

## References

[1] WHO, WHO Coronavirus Disease (COVID-19) Dashboard | WHO Coronavirus Disease (COVID-19) Dashboard, Who. (2020). https://covid19.who.int/ (accessed October 16, 2020).

[2] OPAS, OPAS/OMS | Organização Pan-Americana da Saúde, OPAS. (2020). https://www.paho.org/pt (accessed October 16, 2020).

[3] E. Aromataris, Z. Munn, M. Peters, C. Godfrey, P. McInerney, Z. Munn, A. Tricco, H. Khalil, Chapter 11: Scoping reviews, JBI Rev. Man. (2019). https://doi.org/10.46658/jbirm-20-01.

[4] D. Levac, H. Colquhoun, K.K. O’Brien, Scoping studies: Advancing the methodology, Implement. Sci. 5 (2010) 69. https://doi.org/10.1186/1748-5908-5-69.

[5] J. Strumillo, K.E. Nowak, A. Krokosz, A. Rodacka, M. Puchala, G. Bartosz, The role of resveratrol and melatonin in the nitric oxide and its oxidation products mediated functional and structural modifications of two glycolytic enzymes: GAPDH and LDH, Biochim. Biophys. Acta - Gen. Subj. 1862 (2018) 877–885. https://doi.org/10.1016/j.bbagen.2017.12.017.

[6] A.C. Tricco, E. Lillie, W. Zarin, K.K. O’Brien, H. Colquhoun, D. Levac, D. Moher, M.D.J. Peters, T. Horsley, L. Weeks, S. Hempel, E.A. Akl, C. Chang, J. McGowan, L. Stewart, L. Hartling, A. Aldcroft, M.G. Wilson, C. Garritty, S. Lewin,C.M. Godfrey, M.T. MacDonald, E. V. Langlois, K. Soares-Weiser, J. Moriarty, T. Clifford, Ö. Tunçalp, S.E. Straus, PRISMA extension for scoping reviews (PRISMA-ScR): Checklist and explanation, Ann. Intern. Med. 169 (2018) 467–473. https://doi.org/10.7326/M18-0850.

[7] C. Long, H. Xu, Q. Shen, X. Zhang, B. Fan, C. Wang, B. Zeng, Z. Li, X. Li, H. Li, Diagnosis of the Coronavirus disease (COVID-19): rRT-PCR or CT?, Eur. J. Radiol. 126 (2020) 108961. https://doi.org/10.1016/j.ejrad.2020.108961.

[8] X.T. Ye, Y.L. Luo, S.C. Xia, Q.F. Sun, J.G. Ding, Y. Zhou, W. Chen, X.F. Wang, W.W. Zhang, W.J. Du, Z.W. Ruan, L. Hong, Clinical efficacy of lopinavir/ritonavir in the treatment of Coronavirus disease 2019, Eur. Rev. Med. Pharmacol. Sci. 24 (2020) 3390–3396. https://doi.org/10.26355/eurrev_202003_20706.

[9] R. Liu, H. Han, F. Liu, Z. Lv, K. Wu, Y. Liu, Y. Feng, C. Zhu, Positive rate of RT-PCR detection of SARS-CoV-2 infection in 4880 cases from one hospital in Wuhan, China, from Jan to Feb 2020, Clin. Chim. Acta. 505 (2020) 172–175. https://doi.org/10.1016/j.cca.2020.03.009.

[10] M. Li, W. Peng, M. Chen, Q. Zhu, X. Zou, X. Long, CT imaging features and image evolution characteristics of coronavirus disease 2019, Zhong Nan Da Xue Xue Bao. Yi Xue Ban. 45 (2020) 243–249. https://doi.org/10.11817/j.issn.1672-7347.2020.200168.

[11] B. Cao, Y. Wang, D. Wen, W. Liu, J. Wang, G. Fan, L. Ruan, B. Song, Y. Cai, M. Wei, X. Li, J. Xia, N. Chen, J. Xiang, T. Yu, T. Bai, X. Xie, L. Zhang, C. Li, Y. Yuan, H. Chen, H. Li, H. Huang, S. Tu, F. Gong, Y. Liu, Y. Wei, C. Dong, F. Zhou, X. Gu, J. Xu, Z. Liu, Y. Zhang, H. Li, L. Shang, K. Wang, K. Li, X. Zhou, X. Dong, Z. Qu, S. Lu, X. Hu, S. Ruan, S. Luo, J. Wu, L. Peng, F. Cheng, L. Pan, J. Zou, C. Jia, J. Wang, X. Liu, S. Wang, X. Wu, Q. Ge, J. He, H. Zhan, F. Qiu, L. Guo, C. Huang, T. Jaki, F.G. Hayden, P.W. Horby, D. Zhang, C. Wang, A Trial of Lopinavir–Ritonavir in Adults Hospitalized with Severe Covid-19, N. Engl. J. Med. 382 (2020) 1787–1799. https://doi.org/10.1056/NEJMoa2001282.

[12] Z. Luo, N. Wang, P. Liu, Q. Guo, L. Ran, F. Wang, Y. Tang, Q. Li, Association between chest CT features and clinical course of Coronavirus Disease 2019, Respir. Med. 168 (2020) 105989. https://doi.org/10.1016/j.rmed.2020.105989.

[13] X. Xu, C. Yu, J. Qu, L. Zhang, S. Jiang, D. Huang, B. Chen, Z. Zhang, W. Guan, Z. Ling, R. Jiang, T. Hu, Y. Ding, L. Lin, Q. Gan, L. Luo, X. Tang, J. Liu, Imaging and clinical features of patients with 2019 novel coronavirus SARS-CoV-2, Eur. J. Nucl. Med. Mol. Imaging. 47 (2020) 1275–1280. https://doi.org/10.1007/s00259-020-04735-9.

[14] Y. Jin, M. Wang, Z. Zuo, C. Fan, F. Ye, Z. Cai, Y. Wang, H. Cui, K. Pan, A. Xu, Diagnostic value and dynamic variance of serum antibody in coronavirus disease 2019, Int. J. Infect. Dis. 94 (2020) 49–52. https://doi.org/10.1016/j.ijid.2020.03.065.

[15] N. Ford, M. Vitoria, A. Rangaraj, S.L. Norris, A. Calmy, M. Doherty, Systematic review of the efficacy and safety of antiretroviral drugs against SARS, MERS or COVID-19: initial assessment, J. Int. AIDS Soc. 23 (2020). https://doi.org/10.1002/jia2.25489.

[16] B. Shen, Y. Zheng, X. Zhang, W. Zhang, D. Wang, J. Jin, R. Lin, Y. Zhang, G. Zhu, H. Zhu, J. Li, J. Xu, X. Ding, S. Chen, R. Lu, Z. He, H. Zhao, L. Ying, C. Zhang, D. Lv, B. Chen, J. Chen, J. Zhu, B. Hu, C. Hong, X. Xu, J. Chen, C. Liu, K. Zhou, J. Li,G. Zhao, W. Shen, C. Chen, C. Shao, X. Shen, J. Song, Z. Wang, Y. Meng, C. Wang, J. Han, A. Chen, D. Lu, B. Qian, H. Chen, H. Gao, Clinical evaluation of a rapid colloidal gold immunochromatography assay for SARS-Cov-2 IgM/IgG, Am. J. Transl. Res. 12 (2020) 1348–1354. www.ajtr.org (accessed October 22, 2020).

[17] L. Deng, C. Li, Q. Zeng, X. Liu, X. Li, H. Zhang, Z. Hong, J. Xia, Arbidol combined with LPV/r versus LPV/r alone against Corona Virus Disease 2019: A retrospective cohort study, J. Infect. 81 (2020) e1–e5. https://doi.org/10.1016/j.jinf.2020.03.002.

[18] S. Tabata, M. Ikeda, S. Noguchi, Y. Kitagawa, M. Matsuoka, K. Miyoshi, N. Tarumoto, J. Sakai, T. Ito, S. Maesaki, K. Tamura, T. Maeda, K. Imai, Clinical evaluation of an immunochromatographic IgM/IgG antibody assay and chest computed tomography for the diagnosis of COVID-19, MedRxiv. (2020) 2020.04.22.20075564. https://doi.org/10.1101/2020.04.22.20075564.

[19] T. Klopfenstein, S. Zayet, A. Lohse, J.C. Balblanc, J. Badie, P.Y. Royer, L. Toko, C. Mezher, N.J. Kadiane-Oussou, M. Bossert, A.M. Bozgan, A. Charpentier, M.F. Roux, R. Contreras, I. Mazurier, P. Dussert, V. Gendrin, T. Conrozier, Tocilizumab therapy reduced intensive care unit admissions and/or mortality in COVID-19 patients, Med. Mal. Infect. 50 (2020) 397–400. https://doi.org/10.1016/j.medmal.2020.05.001.

[20] R. Alattar, T.B.H. Ibrahim, S.H. Shaar, S. Abdalla, K. Shukri, J.N. Daghfal, M.Y. Khatib, M. Aboukamar, M. Abukhattab, H.A. Alsoub, M.A. Almaslamani, A.S. Omrani, Tocilizumab for the treatment of severe coronavirus disease 2019, J. Med. Virol. 92 (2020) 2042–2049. https://doi.org/10.1002/jmv.25964.

[21] Y. Wu, T. Wang, C. Guo, D. Zhang, X. Ge, Z. Huang, X. Zhou, Y. Li, Q. Peng, J. Li, Plasminogen improves lung lesions and hypoxemia in patients with COVID-19, QJM. 113 (2020) 539–545. https://doi.org/10.1093/qjmed/hcaa121.

[22] P. Gautret, J.C. Lagier, P. Parola, V.T. Hoang, L. Meddeb, M. Mailhe, B. Doudier, J. Courjon, V. Giordanengo, V.E. Vieira, H. Tissot Dupont, S. Honoré, P. Colson, E. Chabrière, B. La Scola, J.M. Rolain, P. Brouqui, D. Raoult, Hydroxychloroquine and azithromycin as a treatment of COVID-19: results of an open-label non-randomized clinical trial, Int. J. Antimicrob. Agents. 56 (2020) 105949. https://doi.org/10.1016/j.ijantimicag.2020.105949.

[23] L. Li, L. Qin, Z. Xu, Y. Yin, X. Wang, B. Kong, J. Bai, Y. Lu, Z. Fang, Q. Song, K. Cao, D. Liu, G. Wang, Q. Xu, X. Fang, S. Zhang, J. Xia, J. Xia, Artificial Intelligence Distinguishes COVID-19 from Community Acquired Pneumonia on Chest CT, Radiology. 296 (2020) E65–E71. https://doi.org/10.1148/radiol.2020200905.

[24] T. Ai, Z. Yang, H. Hou, C. Zhan, C. Chen, W. Lv, Q. Tao, Z. Sun, L. Xia, Correlation of Chest CT and RT-PCR Testing for Coronavirus Disease 2019 (COVID- 19) in China: A Report of 1014 Cases, Radiology. 296 (2020) E32–E40. https://doi.org/10.1148/radiol.2020200642.

[25] L. Zha, S. Li, L. Pan, B. Tefsen, Y. Li, N. French, L. Chen, G. Yang, E. V. Villanueva, Corticosteroid treatment of patients with coronavirus disease 2019 (COVID-19), Med. J. Aust. 212 (2020) 416–420. https://doi.org/10.5694/mja2.50577.

[26] P. Luo, Y. Liu, L. Qiu, X. Liu, D. Liu, J. Li, Tocilizumab treatment in COVID- 19: A single center experience, J. Med. Virol. 92 (2020) 814–818. https://doi.org/10.1002/jmv.25801.

[27] X. Chen, Y. Tang, Y. Mo, S. Li, D. Lin, Z. Yang, Z. Yang, H. Sun, J. Qiu, Y. Liao, J. Xiao, X. Chen, X. Wu, R. Wu, Z. Dai, A diagnostic model for coronavirus disease 2019 (COVID-19) based on radiological semantic and clinical features: a multi- center study, Eur. Radiol. 30 (2020) 4893–4902. https://doi.org/10.1007/s00330-020-06829-2.

[28] X. Shi, Y. Lu, R. Li, Y. Tang, N. Shi, F. Song, F. Shan, G. Chen, P. Song, Y. Shi, Evaluation of antiviral therapies for coronavirus disease 2019 pneumonia in Shanghai, China, J. Med. Virol. 92 (2020) 1922–1931. https://doi.org/10.1002/jmv.25893.

[29] P. Sarma, H. Kaur, H. Kumar, D. Mahendru, P. Avti, A. Bhattacharyya, M. Prajapat, N. Shekhar, S. Kumar, R. Singh, A. Singh, D.P. Dhibar, A. Prakash, B. Medhi, Virological and clinical cure in COVID-19 patients treated with hydroxychloroquine: A systematic review and meta-analysis, J. Med. Virol. 92 (2020) 776–785. https://doi.org/10.1002/jmv.25898.

[30] Z. Zhu, Z. Lu, T. Xu, C. Chen, G. Yang, T. Zha, J. Lu, Y. Xue, Arbidol monotherapy is superior to lopinavir/ritonavir in treating COVID-19, J. Infect. 81 (2020) e21–e23. https://doi.org/10.1016/j.jinf.2020.03.060.

[31] D. Ferrari, A. Motta, M. Strollo, G. Banfi, M. Locatelli, Routine blood tests as a potential diagnostic tool for COVID-19, Clin. Chem. Lab. Med. 58 (2020) 1095–1099. https://doi.org/10.1515/cclm-2020-0398.

[32] C. Bao, X. Liu, H. Zhang, Y. Li, J. Liu, Coronavirus Disease 2019 (COVID-19) CT Findings: A Systematic Review and Meta-analysis, J. Am. Coll. Radiol. 17 (2020) 701–709. https://doi.org/10.1016/j.jacr.2020.03.006.

[33] Y. Wang, D. Zhang, G. Du, R. Du, J. Zhao, Y. Jin, S. Fu, L. Gao, Z. Cheng, Q. Lu, Y. Hu, G. Luo, K. Wang, Y. Lu, H. Li, S. Wang, S. Ruan, C. Yang, C. Mei, Y. Wang, D. Ding, F. Wu, X. Tang, X. Ye, Y. Ye, B. Liu, J. Yang, W. Yin, A. Wang, G. Fan, F. Zhou, Z. Liu, X. Gu, J. Xu, L. Shang, Y. Zhang, L. Cao, T. Guo, Y. Wan, H. Qin, Y. Jiang, T. Jaki, F.G. Hayden, P.W. Horby, B. Cao, C. Wang, Remdesivir in adults with severe COVID-19: a randomised, double-blind, placebo-controlled, multicentre trial, Lancet. 395 (2020) 1569–1578. https://doi.org/10.1016/S0140-6736(20)31022-9.

[34] F. Liu, Q. Zhang, C. Huang, C. Shi, L. Wang, N. Shi, C. Fang, F. Shan, X. Mei, J. Shi, F. Song, Z. Yang, Z. Ding, X. Su, H. Lu, T. Zhu, Z. Zhang, L. Shi, Y. Shi, CT quantification of pneumonia lesions in early days predicts progression to severe illness in a cohort of COVID-19 patients, Theranostics. 10 (2020) 5613–5622. https://doi.org/10.7150/thno.45985.

[35] L. Gao, J. Zhang, Pulmonary High-Resolution Computed Tomography (HRCT) Findings of Patients with Early-Stage Coronavirus Disease 2019 (COVID-19) in Hangzhou, China, Med. Sci. Monit. 26 (2020) e923885–1. https://doi.org/10.12659/MSM.923885.

[36] R. Mardani, A. Ahmadi Vasmehjani, F. Zali, A. Gholami, S.D. Mousavi Nasab, H. Kaghazian, M. Kaviani, N. Ahmadi, Laboratory Parameters in Detection of COVID- 19 Patients with Positive RT-PCR; a Diagnostic Accuracy Study., Arch. Acad. Emerg. Med. 8 (2020) e43. https://doi.org/10.22037/aaem.v8i1.632.

[37] W. Chen, Y. Lan, X. Yuan, X. Deng, Y. Li, X. Cai, L. Li, R. He, Y. Tan, X. Deng, M. Gao, G. Tang, L. Zhao, J. Wang, Q. Fan, C. Wen, Y. Tong, Y. Tang, F. Hu, F. Li, X. Tang, Detectable 2019-nCoV viral RNA in blood is a strong indicator for the further clinical severity, Emerg. Microbes Infect. 9 (2020) 469–473. https://doi.org/10.1080/22221751.2020.1732837.

[38] W. Wang, Y. Xu, R. Gao, R. Lu, K. Han, G. Wu, W. Tan, Detection of SARS-CoV-2 in Different Types of Clinical Specimens, JAMA - J. Am. Med. Assoc. 323 (2020) 1843–1844. https://doi.org/10.1001/jama.2020.3786.

[39] F. Colavita, D. Lapa, F. Carletti, E. Lalle, L. Bordi, P. Marsella, E. Nicastri, N. Bevilacqua, M.L. Giancola, A. Corpolongo, G. Ippolito, M.R. Capobianchi, C. Castilletti, SARS-CoV-2 Isolation From Ocular Secretions of a Patient With COVID-19 in Italy With Prolonged Viral RNA Detection, Ann. Intern. Med. 173 (2020) 242–243. https://doi.org/10.7326/M20-1176.

[40] K.S. Cheung, I.F.N. Hung, P.P.Y. Chan, K.C. Lung, E. Tso, R. Liu, Y.Y. Ng, M.Y. Chu, T.W.H. Chung, A.R. Tam, C.C.Y. Yip, K.H. Leung, A.Y.F. Fung, R.R. Zhang, Y. Lin, H.M. Cheng, A.J.X. Zhang, K.K.W. To, K.H. Chan, K.Y. Yuen, W.K. Leung, Gastrointestinal Manifestations of SARS-CoV-2 Infection and Virus Load in Fecal Samples From a Hong Kong Cohort: Systematic Review and Meta-analysis, Gastroenterology. 159 (2020) 81–95. https://doi.org/10.1053/j.gastro.2020.03.065.

[41] D. Li, M. Jin, P. Bao, W. Zhao, S. Zhang, Clinical Characteristics and Results of Semen Tests Among Men With Coronavirus Disease 2019, JAMA Netw. Open. 3 (2020) e208292. https://doi.org/10.1001/jamanetworkopen.2020.8292.

[42] F. Yu, L. Yan, N. Wang, S. Yang, L. Wang, Y. Tang, G. Gao, S. Wang, C. Ma,R. Xie, F. Wang, C. Tan, L. Zhu, Y. Guo, F. Zhang, Quantitative Detection and Viral Load Analysis of SARS-CoV-2 in Infected Patients, Clin. Infect. Dis. 71 (2020) 793–798. https://doi.org/10.1093/cid/ciaa345.

[43] D. Xu, F. Zhou, W. Sun, L. Chen, L. Lan, H. Li, F. Xiao, Y. Li, V.B. Kolachalama, Y. Li, X. Wang, H. Xu, Relationship Between serum SARS-CoV-2 nucleic acid(RNAemia) and Organ Damage in COVID-19 Patients: A Cohort Study, Clin. Infect. Dis. (2020). https://doi.org/10.1093/cid/ciaa1085.

[44] K.K.-W. To, I.F.-N. Hung, J.D. Ip, A.W.-H. Chu, W.-M. Chan, A.R. Tam, C.H.- Y. Fong, S. Yuan, H.-W. Tsoi, A.C.-K. Ng, L.L.-Y. Lee, P. Wan, E. Tso, W.-K. To, D. Tsang, K.-H. Chan, J.-D. Huang, K.-H. Kok, V.C.-C. Cheng, K.-Y. Yuen, COVID-19 re-infection by a phylogenetically distinct SARS-coronavirus-2 strain confirmed by whole genome sequencing, Clin. Infect. Dis. (2020). https://doi.org/10.1093/cid/ciaa1275.

[45] J. Van Elslande, P. Vermeersch, K. Vandervoort, T. Wawina-Bokalanga, B. Vanmechelen, E. Wollants, L. Laenen, E. André, M. Van Ranst, K. Lagrou, P. Maes, Symptomatic SARS-CoV-2 reinfection by a phylogenetically distinct strain, Clin. Infect. Dis. (2020). https://doi.org/10.1093/cid/ciaa1330.

[46] M. Lv, X. Luo, J. Estill, Y. Liu, M. Ren, J. Wang, Q. Wang, S. Zhao, X. Wang, S. Yang, X. Feng, W. Li, E. Liu, X. Zhang, L. Wang, Q. Zhou, W. Meng, X. Qi, Y. Xun, X. Yu, Y. Chen, X. Liu, N. Yang, S. Lu, P. Du, Y. Ma, Z. Wang, Q. Shi, H. Zhang, Q. Guo, Y. Yang, B. Yang, S. Wu, X. Wang, Coronavirus disease (COVID-19): A scoping review, Eurosurveillance. 25 (2020). https://doi.org/10.2807/1560-7917.ES.2020.25.15.2000125.

[47] W. Wang, Y. Xu, R. Gao, R. Lu, K. Han, G. Wu, W. Tan, Detection of SARS-CoV-2 in Different Types of Clinical Specimens, JAMA - J. Am. Med. Assoc. 323 (2020) 1843–1844. https://doi.org/10.1001/jama.2020.3786.

[48] X. Yao, F. Ye, M. Zhang, C. Cui, B. Huang, P. Niu, X. Liu, L. Zhao, E. Dong, C. Song, S. Zhan, R. Lu, H. Li, W. Tan, D. Liu, In Vitro Antiviral Activity and Projection of Optimized Dosing Design of Hydroxychloroquine for the Treatment of Severe Acute Respiratory Syndrome Coronavirus 2 (SARS-CoV-2), Clin. Infect. Dis. 71 (2020) 732–739. https://doi.org/10.1093/cid/ciaa237.

[49] J. Chen, D. Liu, L. Liu, P. Liu, Q. Xu, L. Xia, Y. Ling, D. Huang, S. Song, D. Zhang, Z. Qian, T. Li, Y. Shen, H. Lu, A pilot study of hydroxychloroquine in treatment of patients with moderate COVID-19, Zhejiang Da Xue Xue Bao. Yi Xue Ban. 49 (2020) 215–219. https://doi.org/10.3785/j.issn.1008-9292.2020.03.03.

[50] P. Gautret, J.C. Lagier, P. Parola, V.T. Hoang, L. Meddeb, M. Mailhe, B. Doudier, J. Courjon, V. Giordanengo, V.E. Vieira, H. Tissot Dupont, S. Honoré, P. Colson, E. Chabrière, B. La Scola, J.M. Rolain, P. Brouqui, D. Raoult, Hydroxychloroquine and azithromycin as a treatment of COVID-19: results of an open-label non-randomized clinical trial, Int. J. Antimicrob. Agents. 56 (2020). https://doi.org/10.1016/j.ijantimicag.2020.105949.

[51] J. Gao, Z. Tian, X. Yang, Breakthrough: Chloroquine phosphate has shown apparent efficacy in treatment of COVID-19 associated pneumonia in clinical studies, Biosci. Trends. 14 (2020). https://doi.org/10.5582/BST.2020.01047.

[52] J.M. Sanders, M.L. Monogue, T.Z. Jodlowski, J.B. Cutrell, Pharmacologic Treatments for Coronavirus Disease 2019 (COVID-19): A Review, JAMA - J. Am. Med. Assoc. 323 (2020) 1824–1836. https://doi.org/10.1001/jama.2020.6019.

[53] P. Toniati, S. Piva, M. Cattalini, E. Garrafa, F. Regola, F. Castelli, F. Franceschini, P. Airò, C. Bazzani, E.A. Beindorf, M. Berlendis, M. Bezzi, N. Bossini, M. Castellano, S. Cattaneo, I. Cavazzana, G.B. Contessi, M. Crippa, A. Delbarba, E. De Peri, A. Faletti, M. Filippini, M. Frassi, M. Gaggiotti, R. Gorla, M. Lanspa, S. Lorenzotti, R. Marino, R. Maroldi, M. Metra, A. Matteelli, D. Modina, G. Moioli, G. Montani, M.L. Muiesan, S. Odolini, E. Peli, S. Pesenti, M.C. Pezzoli, I. Pirola, A. Pozzi, A. Proto, F.A. Rasulo, G. Renisi, C. Ricci, D. Rizzoni, G. Romanelli, M. Rossi, M. Salvetti, F. Scolari, L. Signorini, M. Taglietti, G. Tomasoni, L.R. Tomasoni, F. Turla, A. Valsecchi, D. Zani, F. Zuccalà, F. Zunica, E. Focà, L. Andreoli, N. Latronico, Tocilizumab for the treatment of severe COVID-19 pneumonia with hyperinflammatory syndrome and acute respiratory failure: A single center study of 100 patients in Brescia, Italy, Autoimmun. Rev. 19 (2020) 102568. https://doi.org/10.1016/j.autrev.2020.102568.

[54] C. Campochiaro, E. Della-Torre, G. Cavalli, G. De Luca, M. Ripa, N. Boffini, A. Tomelleri, E. Baldissera, P. Rovere-Querini, A. Ruggeri, G. Monti, F. De Cobelli, A. Zangrillo, M. Tresoldi, A. Castagna, L. Dagna, Efficacy and safety of tocilizumab in severe COVID-19 patients: a single-centre retrospective cohort study, Eur. J. Intern. Med. 76 (2020) 43–49. https://doi.org/10.1016/j.ejim.2020.05.021.

[55] H. Pan, R. Peto, Q.A. Karim, M. Alejandria, A.M. Henao-Restrepo, C.H. García, et al. Repurposed antiviral drugs for COVID-19-interim WHO SOLIDARITY trial results, Pre-print, medRxiV. https://doi.org/10.1101/2020.10.15.20209817

[56] A. Saghazadeh, N. Rezaei, Towards treatment planning of COVID-19: Rationale and hypothesis for the use of multiple immunosuppressive agents: Anti-antibodies, immunoglobulins, and corticosteroids, Int. Immunopharmacol. 84 (2020). https://doi.org/10.1016/j.intimp.2020.106560.

